# Genetic correlation analysis identifies *TMEM106B, ACE*, and *ERC2* as genetic loci shared between Alzheimer’s disease and primary psychiatric disorders

**DOI:** 10.1101/2025.10.03.25336901

**Authors:** Ajneesh Kumar, Nicholas R. Ray, Jiji T. Kurup, Pamela Del Rosario, Masood Manoochehri, Colin Stein, Alyssa N. De Vito, Brenna Cholerton, Robert A. Sweet, Phil L. de Jager, Hans-Ulrich Klein, Michael L. Cuccaro, Gary W. Beecham, Edward D. Huey, Christiane Reitz

## Abstract

**Background:** Neuropsychiatric symptoms (NPS) occur in up to 85% of Alzheimer’s disease (AD) cases. Current treatments—repurposed from psychiatric disorders despite limited understanding of etiologic overlap—are often ineffective.

**Methods:** To characterize the genetic overlap between AD and major psychiatric disorders and identify shared molecular pathways we conducted genetic correlation analyses between AD and depression, schizophrenia, bipolar disorder and anxiety using MiXeR and LAVA using GWAS summary statistics (AD: n=487,511; bipolar disorder: n=413,466; depression: n=1,154,267; schizophrenia: n=130,644; anxiety: n=1,096,458).

**Results:** Genetic correlation analyses followed by fine mapping and functional analyses identified a missense variant in *TMEM106B* (rs3173615) shared between AD and depression and anxiety, a regulatory region variant in *ACE* (rs4292) shared between AD/schizophrenia, and two NMD transcript variants in *ERC2* (rs17288728; rs815460) shared between AD/anxiety.

**Conclusion:** The specific molecular pathways associated with these variants provide critical information on shared etiologic components underlying these traits, and inform development of improved therapeutic targets.

## 1. INTRODUCTION

As the seventh leading cause of death in the US, over six million affected individuals in the U.S alone, and incalculable costs in terms of human suffering, Alzheimer’s disease (AD) is a high priority for the development of effective, safe and affordable therapies.[1] While cognitive impairment is the predominant and often first noticeable symptom, approximately 85% of AD patients will also develop a neuropsychiatric symptom (NPS; e.g., aggression, psychosis, anxiety, apathy, depression, agitation, sleep disturbances, repetitive thoughts and behaviors) at some point in the illness.[2, 3] NPS are associated with greatly decreased quality of life for patients,[4] accelerated disease progression,[5] increased mortality,[5] increased out of home placement,[6] and greatly increased costs of care.[3, 7] In addition, NPS – especially aggression, psychosis, agitation, and sleep disturbances – have a profound effect on caregiver stress, burden, and depressive symptoms, even higher than that of cognitive or functional impairment.[8-11] As such, any mitigation of NPS in AD would be a substantial improvement in quality of life for both patients and their caregivers.

Despite this urgency, our understanding of the etiology of NPS in AD is still limited, and currently available treatments for NPS are often ineffective and associated with serious adverse effects. Pharmacological treatment regimens have generally been borrowed from their indications for psychiatric illness, despite limited understanding to what extent the molecular and genetic architectures underlying AD-associated NPS overlap with primary psychiatric disorders. Indeed, there is evidence that etiology, presentation, course, and treatment responsiveness of NPS in AD and primary psychiatric disorders are at least partly dissimilar. For example, psychosis in AD is much more likely to present with simple delusions related to memory symptoms than the complex delusional systems and auditory hallucinations characteristic of schizophrenia, and has different genetic associations than schizophrenia.[12]

Identification of disease-associated genetic variants, genes and loci can directly point to underlying mechanistic pathways and druggable targets. To start characterizing the genetic overlap between AD and major psychiatric disorders and identify potentially shared causative genetic variants, genes and biological processes we conducted global and local genetic correlation analyses between AD and bipolar disorder, depression, schizophrenia and anxiety followed by functional finemapping, capitalizing on the largest most recent genomic data currently available for these traits.

## 2. METHODS

### 2.1 GWAS Summary Statistics Data Acquisition and Preprocessing

We retrieved the most recent and latest AD and NPS trait (depression, bipolar disorder, anxiety disorder and schizophrenia) GWAS summary statistics; (AD; 85,934 cases, 401,577 controls) [13], depression (340,591 cases, 813,676 controls)[14], bipolar disorder (41,917 cases, 371,549 controls) [15], schizophrenia (53,386 cases, 77,258 controls) [16], and anxiety (1,096,458 total number of samples; Table1) [17]. To maximize sample size and minimize confounding effects of dissimilar ancestries, this study focused on non-Hispanic Whites. To yield similar genome builds across datasets, genomic coordinates for the AD data were lifted down from hg38 to hg37, employing the LiftOver tool from the UCSC Genome Browser (https://genome.ucsc.edu/cgi-bin/hgLiftOver). We then reformatted the summary statistics of AD and all NPS traits by employing the standard data cleaning and preprocessing script from the LDSc tool [18] and subsequently employed LDSc to determine sample overlap by calculating the genetic covariance intercepts between AD and each individual NPS trait.

### 2.2 Global genetic correlation analysis

Capitalizing on these GWAS summary statistics, we employed MiXeR [19] to estimate heritability and polygenicity of each individual phenotype and assess the global genetic correlation between AD and each NPS trait. For a pair of phenotypes, MiXeR quantifies polygenic overlap irrespective of genetic correlation between traits by estimating the total number of shared and trait-specific causal variants (i.e., variants with nonzero additive genetic effect on a trait), visualizing results with Venn diagrams. MiXeR inference of parameters is based on maximizing the log-likelihood function observing a set of GWAS summary statistics given model parameters, while accounting for the LD structure among SNPs and their allele frequencies. In addition, MiXeR estimates genome-wide correlations across all variants (r_g_) along with the correlation of effect sizes within the shared genetic component (r_gs_). Univariate MiXeR analyses for each phenotype were employed to estimate the SNP-based heritability (h^2^_SNP_) and polygenicity (number of variants accounting for 90% of SNP heritability). For each trait, quantile-quantile (Q-Q) plots were constructed for observed versus predicted GWAS p values and partitioned by minor allele frequency and linkage disequilibrium (LD), providing information whether the GWASs were sufficiently powered. We evaluated model fit both for univariate and bivariate analyses (i.e. the ability of the MiXeR model to accurately predict the actual GWAS data) employing modeled versus actual conditional Q-Q plots, negative log-likelihood plots, the Akaike information criterion (AIC) and the Bayesian information criterion (BIC). The conditional Q-Q plots show observed versus expected −log10 p-values in the primary trait as a function of the significance of association with a secondary trait at the level of p□≤□0.1, p□≤ □0.01, and p□≤□0.001, with successive inflated deflection of SNP strata with higher significance in the secondary (conditional) trait indicating cross-trait enrichment. Model fit is demonstrated in these conditional Q-Q plots if the data Q-Q plots (solid lines) are closely reproduced by the model predictions (dashed lines) across all p-value strata. Negative log-likelihood plots visualize the performance of the best model versus models with minimum and maximum polygenic overlap. The minimum model is represented by the point furthest to the left, the maximum model is represented by the point furthest to the right, and the lowest point on the curve (y-axis) indicates better model fit. Support for the MiXeR model is a clearly defined minimum on the negative log-likelihood curve. LD structure was estimated using the 1000 Genomes Phase 3 genotype reference panel [20].

### 2.3 Local genetic correlation analysis

Local genetic correlation analyses between AD and depression, schizophrenia, bipolar disorder, and anxiety were conducted using Local Analysis of [co]Variant Annotation (LAVA) [21]. LAVA is an integrated framework for local *r*_g_ (regression) analysis which can analyze binary as well as continuous phenotypes with varying degrees of sample overlap, and in addition to testing the standard bivariate local *r*_g_’s between two traits can evaluate the local heritability for all traits of interest, and analyze conditional genetic relations between several traits using partial correlation or multiple regression [21]. To calculate local genetic correlation, we utilized the default LD block file provided by LAVA for European populations (https://github.com/cadeleeuw/lava-partitioning) which is based on the 1000Genomes EUR reference population setting the minimum block size to 2500, yielding 2,495 partitioned LD blocks, followed by calculation of bivariate associations. Genetic loci identified by local genetic correlation analysis meeting a significance threshold of P-value < 5 *10^-3 were further visualized by local association plots using LOCUSZOOM [22] The loci that exhibited significant signal for both traits at a genome-wide significant p-value threshold of 5*10^^-5^ in the same region were chosen as the top loci on the basis of local association plots visualization.

### 2.4 Fine mapping analysis and functional follow-up

To conduct functionally informed finemapping of identified loci shared between AD and NPS traits, we utilized the Polyfun+Susie pipeline [23]. PolyFun is a framework to improve fine-mapping accuracy by leveraging genome-wide functional data for a broad set of coding, conserved, regulatory and LD-related annotations from the baseline-LF 2.2.UKB model using an L2-regularized extension of S-LDSC [23-25], and prioritizing variants in enriched functional annotations by specifying prior causal probabilities for fine-mapping methods such as SuSiE or FINEMAP [26, 27]. Variants and loci identified in these analyses were further scrutinized employing ADSP R4 whole-genome sequencing data, information from the AGORA portal (https://agora.adknowledgeportal.org/) [28], multi-omic data from ONTIME (https://ontime.wustl.edu/), methylation and histone modification data from the ROSMAP study [29], HI-C long-range data from the 3D-genome browser (https://3dgenome.fsm.northwestern.edu/tutorial.html) [30], and eQTL data from GTEX [31].

### 2.5 Brain DNA methylation analyses in the ROS/MAP cohort

DNA methylation data came from frozen dorsolateral prefrontal cortex from 761 participants in ROS/MAP[32]. All ROS/MAP participants enroll without known dementia, agree to annual clinical evaluation and agree to brain donation at the time of death. Details of the data generation have been previously published in detail; methylation profiles were generated using the Illumina HumanMethylation450 beadset.[33, 34]. For the present analyses, the β-values reported by the Illumina platform for each probe ranging from 0 (no methylation) to 1 (100% methylation) were utilized as the methylation level measurement for the targeted CG site in a given sample, and a linear regression model was applied to examine identified top loci for differentially methylated regions associated with AD pathology adjusting for age at death, sex, experimental batch, and bisulfite conversion efficiency. β-amyloid load and PHF-tau tangle density were generated as previously described [35, 36].

### 2.6 3D genome organization and long-range chromatin interactions

3D genome organization and long-range chromatin interactions for top loci resulting from the functional finemapping analysis were retrieved employing the 3D genome browser [30] which can simultaneously query and supplement chromatin interaction data with thousands of genetic, epigenetic, and phenotypic datasets, including ChIP-Seq and RNA-Seq data from the ENCODE and Roadmap Epigenomics projects.

### 2.7 eQTL data

eQTL expression data for top loci resulting from the functional fine-mapping analysis were retrieved by querying the GTEx portal (https://www.gtexportal.org/home/).

## 3. RESULTS

### 3.1 Shared global genetic architecture between AD and NPS traits

The results from the univariate MiXeR results are summarized in Supplementary Table 1. SNP-based heritability differed between AD and the four psychiatric traits, with schizophrenia showing the highest heritability (h2 SNP = 0.44 ± 0.06), and depression (h2 SNP = 0.03 ± 0.008), bipolar disorder (h2 SNP = 0.08 ± 0.02), anxiety (h2 SNP = 0.02 ± 0.006) and AD (h2 SNP = 0.08 ± 0.27) showing a lower, comparable heritability. The univariate AIC were all positive (see Supplementary Table 1) indicating sufficient model fit, and univariate Q-Q plots suggested that MiXeR-based predictions provide accurate estimates of the data plots (Supplemental Figure 1).

Quantification of global polygenic overlap using MiXeR identified both unique genetic components underlying each trait, as well as polygenic components shared between AD and each of the four psychiatric traits. At 90% of SNP-heritability explained for each phenotype, MiXeR estimated that ∼1.8k variants causally influence AD, ∼15.3k variants influence depression, ∼15.1k variants influence schizophrenia, ∼15.6k variants influence bipolar disorder, and ∼15.1k variants influence anxiety (Supplemental Table 1; Figure 1). Among these variants, 0.9k variants are shared between AD and depression (representing 50.0% of the genetic variants influencing AD and 5.7% of the variants underlying depression), 1.3k variants are shared between AD and schizophrenia (representing 72.2% of the genetic variants influencing AD and 8.6% of the variants underlying schizophrenia), 1.3k variants are shared between AD and bipolar disorder (representing 72.2% of the genetic variants influencing AD and 8.3% of the variants underlying bipolar disorder), and 1.1k variants are shared between AD and anxiety (representing 61.1% of the genetic variant influencing AD and 7.8% of the underlying anxiety) suggesting significant pleiotropy and genetic correlation of AD with each of the four psychiatric traits. Overall genome-wide genetic correlation (r_g_) with depression was estimated at r_g_= −0.01, schizophrenia at r_g_ = 0.03, bipolar disorder at r_g_ = 0.04, and anxiety at r_g_ = 0.02 (Supplementary Table 2; Figure 1), with correlation of the effect sizes of shared causal variants estimated at ρβ□=□-0.13 (SD□=□0.81) for depression, ρβ□=□0.19 (SD□=□0.62) for schizophrenia, ρβ□=□0.23(SD□=□0.62) for bipolar disorder, and ρβL=L-0.05(SD□=□0.76) for anxiety, and proportion of shared causal variants with concordant effects estimated at 0.45 for depression, 0.57 for schizophrenia, 0.59 for bipolar disorder and 0.44 for anxiety (Supplementary Table 2).

**Figure 1.**
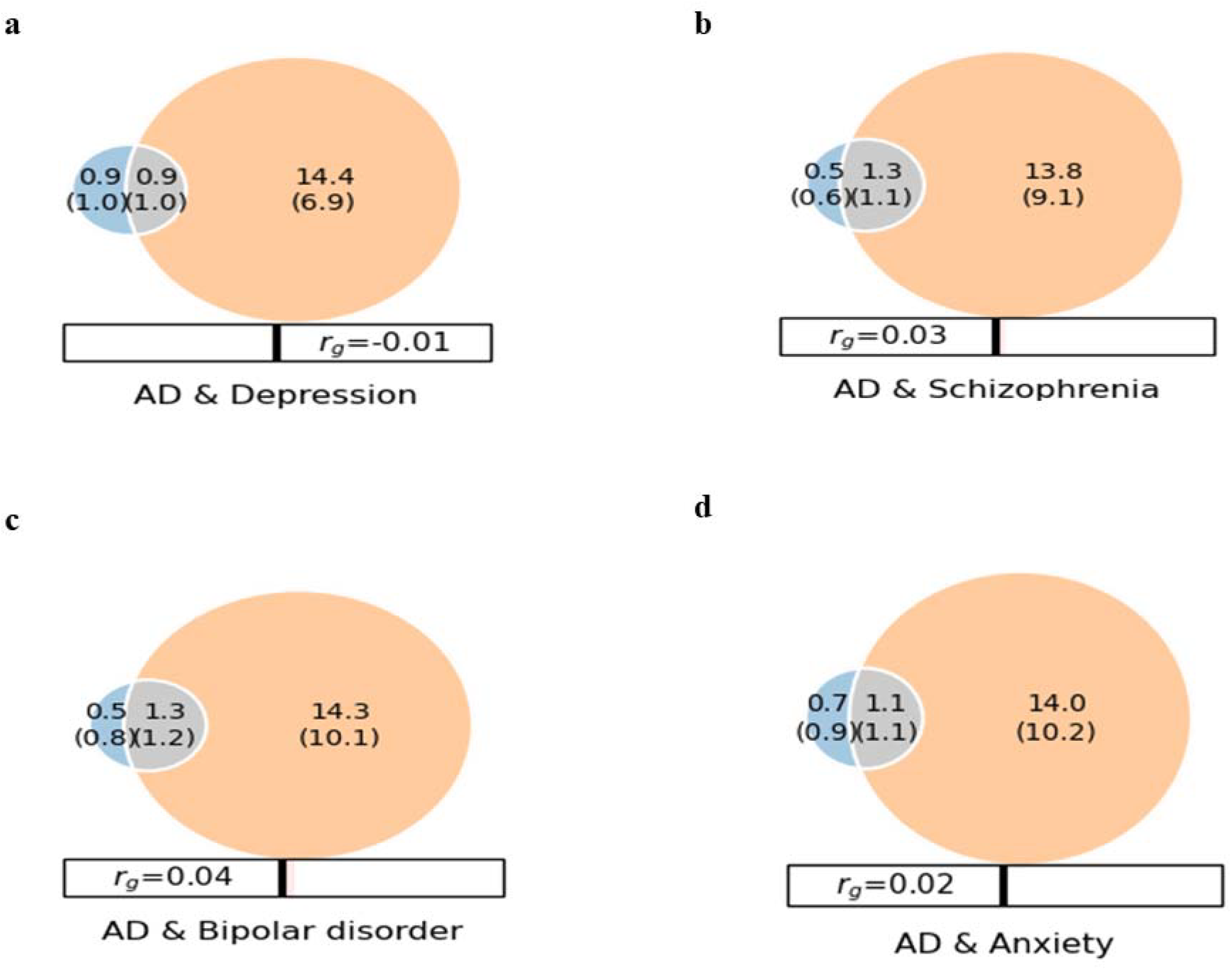
Genome-wide bivariate genetic overlap between Alzheimer’s Disease (AD), depression, schizophrenia, bipolar disorder and anxiety (estimated employing MiXeR), The Venn diagrams display the number of shared and causal genetic variants (in thousands) explaining 90% of SNP heritability in each phenotype, followed by the standard error. The size of the circles reflects the degree of polygenicity. The blue circles represent unique trait-specific variants underlying AD, the orange circles represent unique trait-specific variants underlying the respective psychiatric trait, the grey circles represent shared genetic variants between AD and the respective psychiatric trait. r_g_; Global genetic correlation. (a) AD & Depression, (b) AD & Schizophrenia, (c) AD & Bipolar Disorder, and (d) AD & Anxiety

### 3.2 Local genetic covariance analysis

Local genetic covariance analyses employing LAVA [21], identified 17 genetic loci shared between AD and depression, 23 loci shared between AD and schizophrenia, 17 loci shared between AD and bipolar disorder, and 9 loci shared between AD and anxiety at a significance threshold of P <= 5* 10^-3 (Supplementary Table 3). From these, 14 individual loci met a GWAS threshold of P < 5* 10^-5 individually in both AD and the respective NPS trait and were further assessed using local association analyses, finemapping, and *in silico* functional analyses. Four loci located at 2q37.1, 6p22, 7p21, and 16p13, harboring *ARL4C, ID4, TMEM106B*, and *RBFOX1*, respectively showed genetic covariance with depression, four loci (3q21, 17q21, 17q23, 20q13) harboring *SLC12A8, MAPT/KANSL1, ACE* and *LIME1/STMN3* showed genetic covariance with schizophrenia, and two loci (3p22 and 18q21 harboring *TRANK1* and *ZCCHC2*) showed covariance with bipolar disorder (Table 2). Notably, one locus (20q18) harboring *KCNG1* showed genetic covariance with both schizophrenia and bipolar disorder. Three loci (3p14, 6p21.33, 7p21) harboring *ERC2, MSH5*, and *TMEM106B* showed genetic covariance with anxiety (Table 2).

**Table 1.**
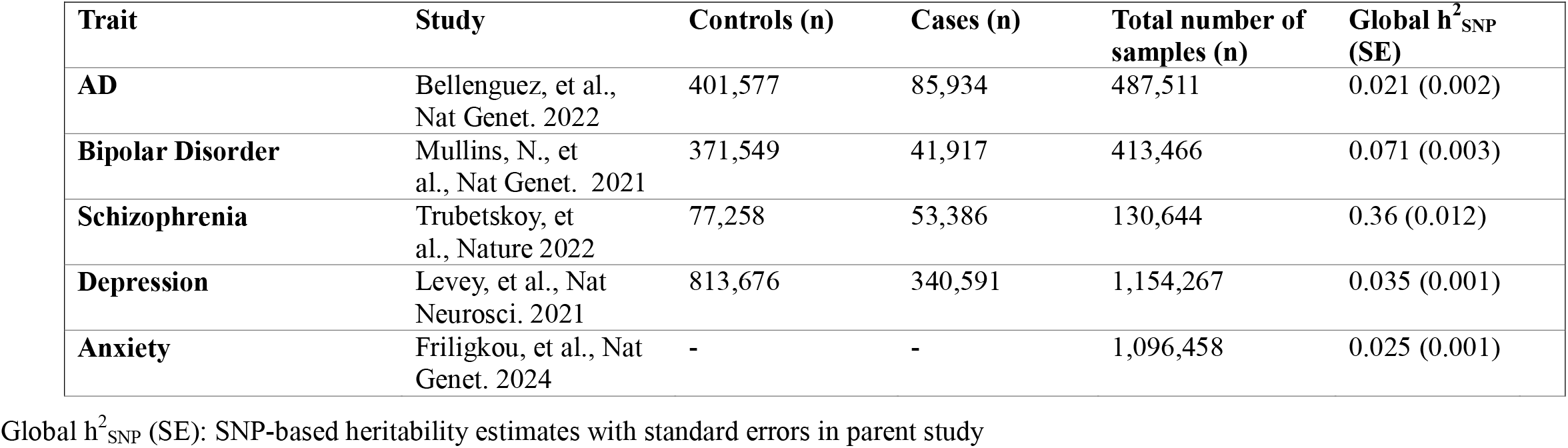
Overview of data utilized in global and local genetic covariance analyses.

| Trait | Study | Controls (n) | Cases (n) | Total number of samples (n) | Global $h^2_{\text{SNP}}$ (SE) |
| --- | --- | --- | --- | --- | --- |
| AD | Bellenguez, et al., Nat Genet. 2022 | 401,577 | 85,934 | 487,511 | 0.021 (0.002) |
| Bipolar Disorder | Mullins, N., et al., Nat Genet. 2021 | 371,549 | 41,917 | 413,466 | 0.071 (0.003) |
| Schizophrenia | Trubetskoy, et al., Nature 2022 | 77,258 | 53,386 | 130,644 | 0.36 (0.012) |
| Depression | Levey, et al., Nat Neurosci. 2021 | 813,676 | 340,591 | 1,154,267 | 0.035 (0.001) |
| Anxiety | Friligkou, et al., Nat Genet. 2024 | - | - | 1,096,458 | 0.025 (0.001) |

Global $h^2_{\text{SNP}}$ (SE): SNP-based heritability estimates with standard errors in parent study

**Table 2.**
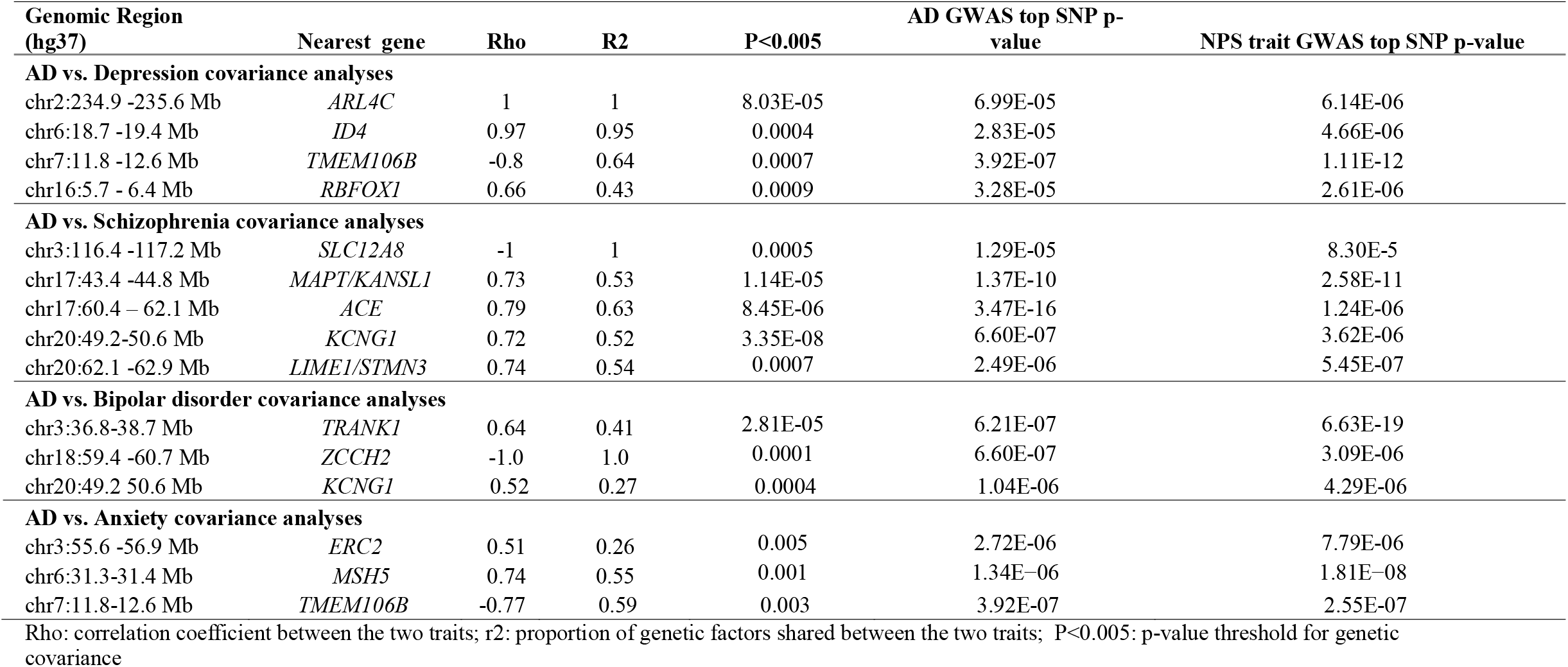
Loci identified in local covariance analyses harboring variants with minimum p-value of 10-5 in both AD and the respective NPS trait summary statistics.

Closer examination of the local association and LD patterns in these regions demonstrated perfect genetic overlap of AD and the respective neuropsychiatric trait association patterns at the 3p14, 7p21, 17q23, and 17q21 loci with strong genetic association peaks for both AD and the respective neuropsychiatric trait localized within the same haplotype blocks within *TMEM106B* (AD/depression/anxiety), *ACE* (AD/schizophrenia), *MAPT/KANSL1* (AD/schizophrenia) and *ERC2* (AD/anxiety) (Figure 2a-d). In addition, local association analyses demonstrated overlapping genetic correlation localized to the same haplotype block within the *KCNG1* locus at 20q18, showing genetic covariance with both schizophrenia and bipolar disorder, albeit with a slightly weaker signal in AD (Figure 2e). The local association plots for the remaining loci are displayed in Supplemental Figures 2-5. *TMEM106B, ACE, MAPT/KANSL1, KCNG1*, and *ERC2* are highly expressed across brain regions (Supplemental Figure 6).

**Figure 2.**
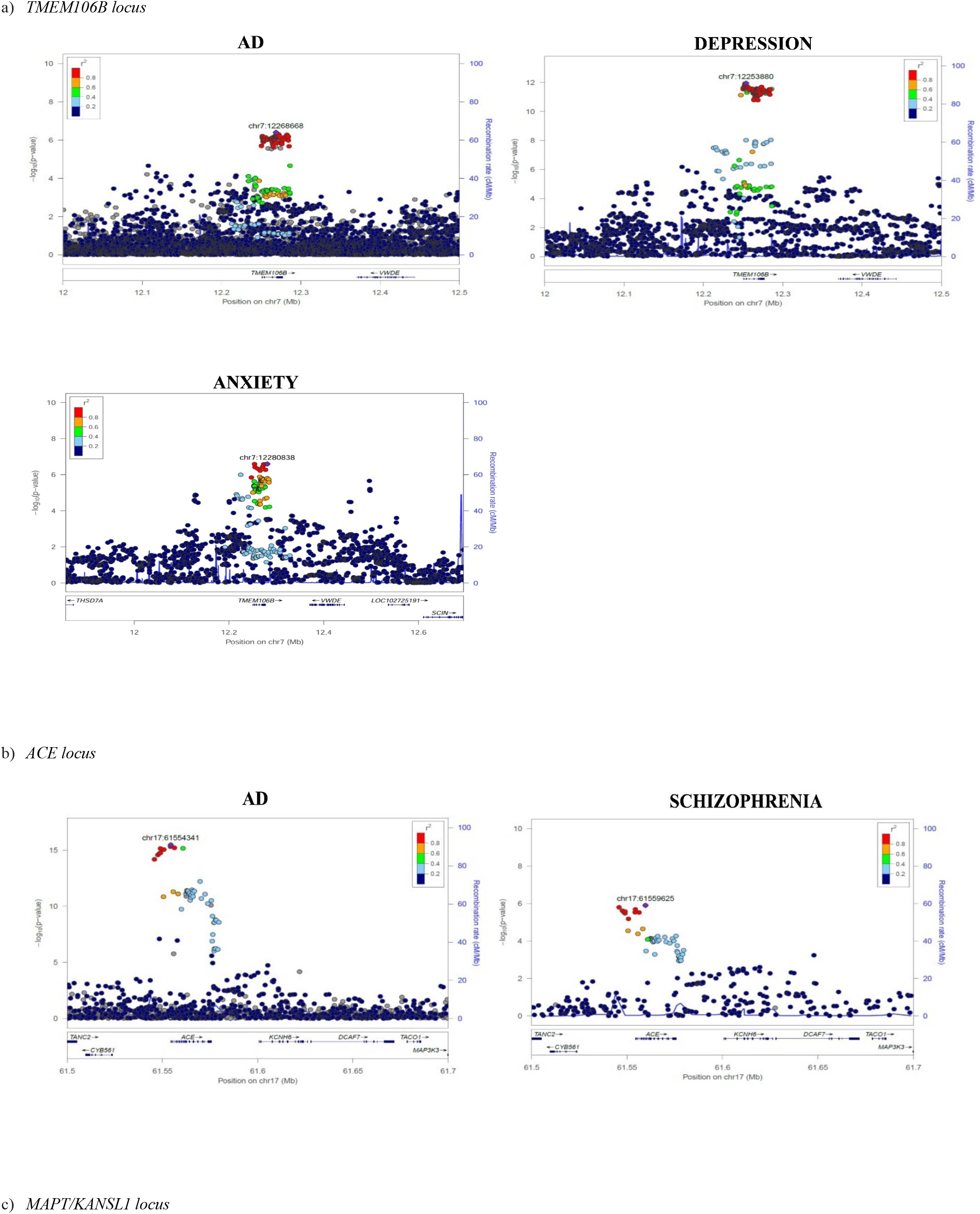
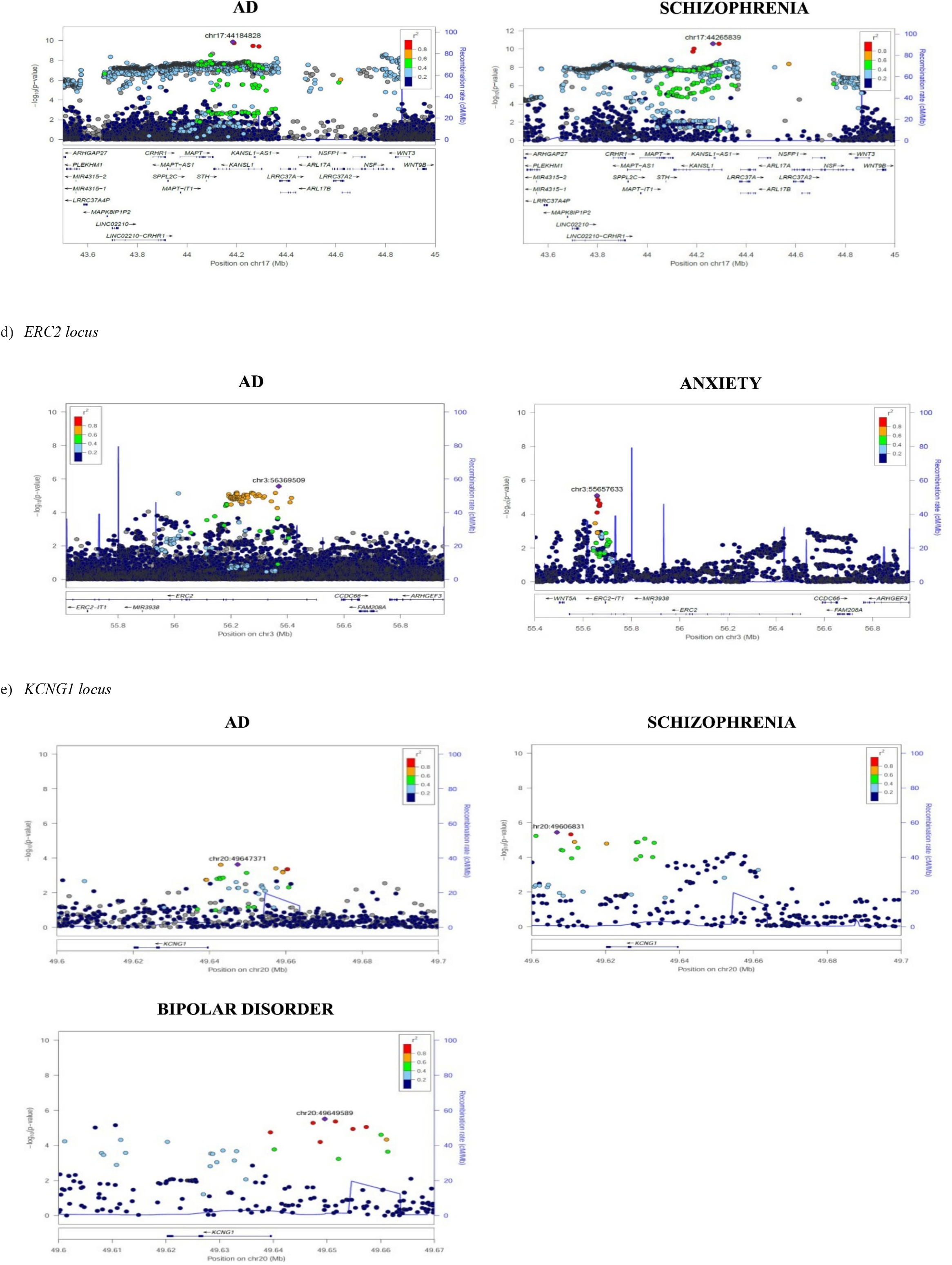
Local association plots for *TMEM106B, ACE, MAPT/KANSL1, KCNG1, ERC2* loci. The x-axis represents the base pair position on the chromosome. The left y-axis shows −log10(p-values) for genetic association, while the right y-axis depicts recombination rates (cM/Mb). The color of the points indicates the linkage disequilibrium (r^2^) with the lead SNP. The left-hand plots show the local association plot for AD, and the right-hand plots show the local association plots for the respective NPS traits analyzed

### 3.3 Functionally informed finemapping of identified top loci

Functionally informed finemapping of the top loci showing genetic covariance between AD and NPS traits using the Polyfun+Susie pipeline [23] identified a credible set harboring a conserved missense variant (rs3173615) at the *TMEM106B* locus (PIP _Depression_ = 0.52; PIP_AD_ = 0.38; PIP_Anxiety_ = 0.79; Figure 3a). rs3173615 has previously been reported as a GWAS top variant associated with hippocampal sclerosis, as well as FTLD/TDP-43[37], and is in high LD with a 3’ UTR 322bp in/del polymorphism. Functionally informed finemapping also identified a credible set harboring a conserved regulatory region variant (rs4292) at the *ACE* locus (PIP_AD_ = 0.78 and PIP _Schizophrenia_ = 0.62; Figure 3b), and two credible sets containing individual NMD transcript variants in the AD haplotype (rs17288728; PIP_AD_ = 0.25) and the anxiety haplotype (rs815460; PIP _Anxiety_ = 0.55) at the *ERC2* locus (Figure 3c). Based on AGORA, *TMEM106B* has a target risk AD score of 1.98 and a genetic AD risk score of 1.82, *ACE* has a AD target risk score of 3.23 and a genetic risk AD score of 2.37, and *ERC2* has a target risk AD score of 4.0, a genetic risk score of 2.04 and a multi-omic score of 1.95. Analysis of proteomic, metabolomic and brain amyloid imaging data from the Washington University ONTIME server demonstrated that rs3173615 is a significant pQTL for *TMEM106B* in both non-Hispanic Whites and individuals of African ancestry and also modulates Histone H2A, while rs4292 is a significant pQTL for *ACE* and is also associated with amyloid burden on brain amyloid imaging (Supplemental Figure 7). In line with this notion, in brain methylation data from the ROS/MAP cohort methylation patterns in *TMEM1056B, ACE* and *ERC2* were associated with amyloid pathology (Supplementary Table 4).

**Figure 3.**
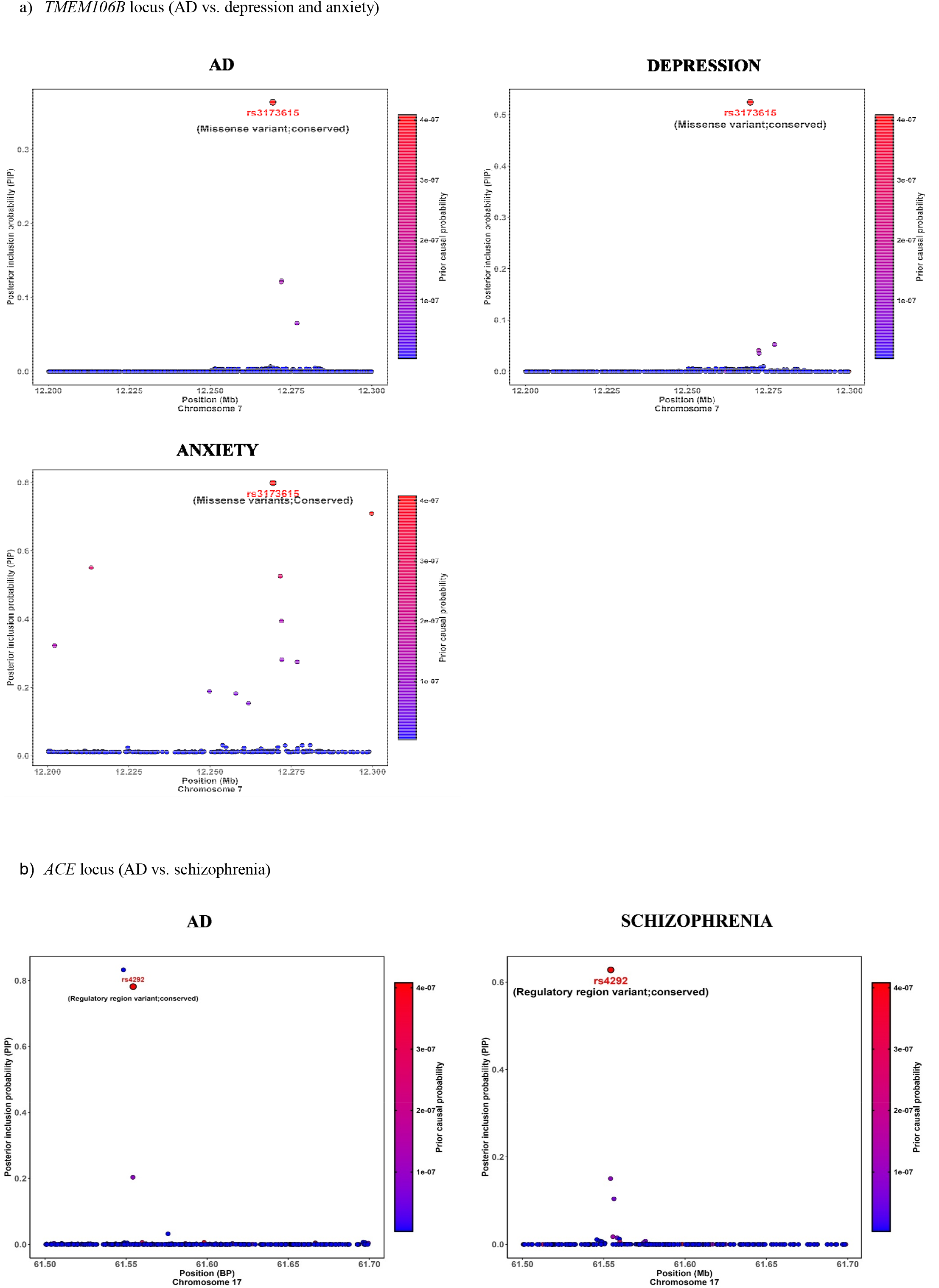
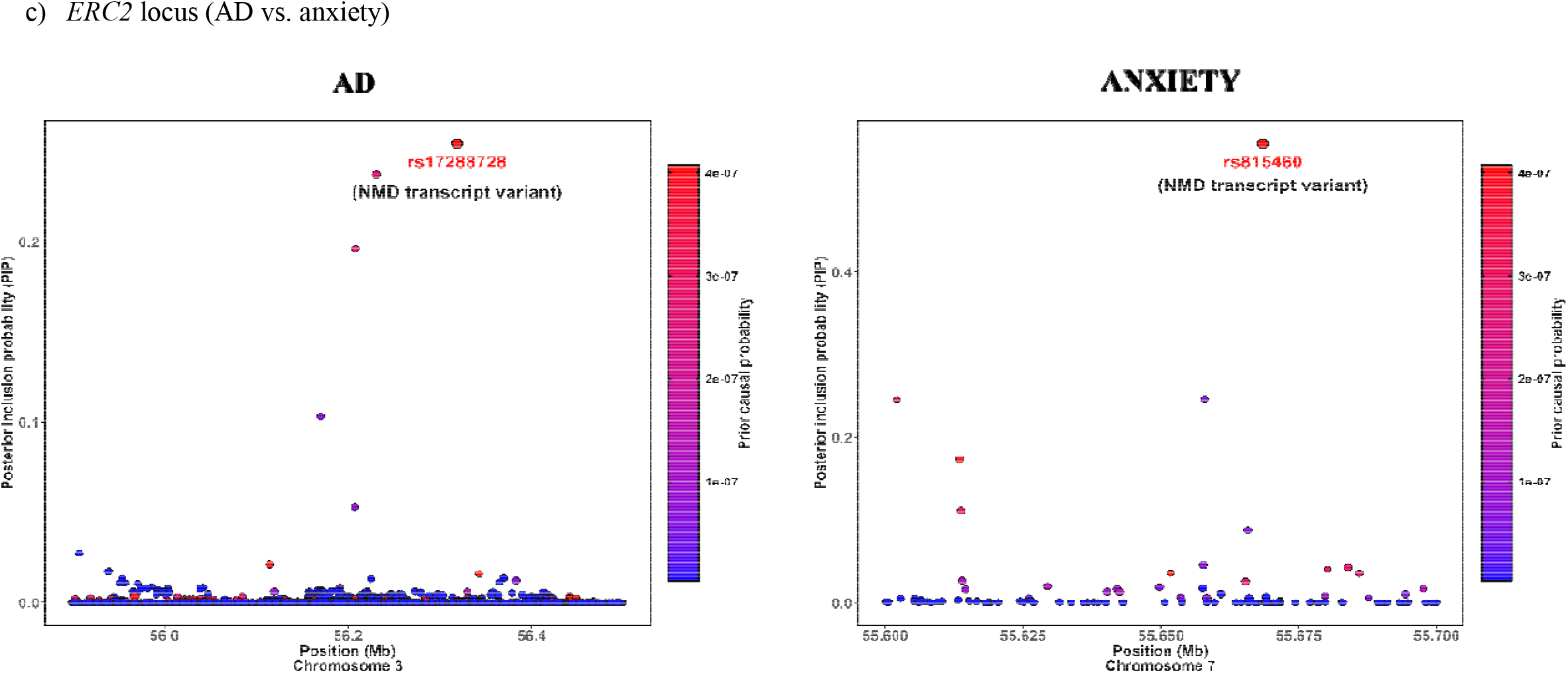
Causal credible sets (≤10 variants) with cumulative PIP > 0.95 identified by functional fine mapping at the *TMEM106B, ACE*, and *ERC2* loci. The x-axis represents the base pair position, the left y-axis shows the PIP values, and the right y-axis shows the posterior causal probability. Each dot represents a genetic variant, with the red dots highlighting variants with the highest PIP value in the credible set

### 3.4 Validation of identified top variants in ADSP R4 sequence data

Analysis of these variants prioritized by fnemapping in ADSP R4 sequence data replicated both the regulatory region variant in *ACE* (rs4292; CADD score: 17.8) as well as the *ERC2* NMD transcript variant rs815460 (CADD score: 19.0) as associated with AD (rs4292; total number of alleles in cases = 7910; total minor allele frequency count in cases = 2835; total number of alleles in controls= 4394; total minor allele frequency count in controls = 1679; χ^*2*^ = 6.73; *P* = 9×10 ^-3^; rs815460; total number of alleles in cases = 8020; total minor allele frequency count in cases = 2924; total number of alleles in controls= 4426; total minor allele frequency count in controls = 1460; χ^*2*^ = 14.91; *P* = 1×10 ^-4^). Analyses of HI-C chromatin interaction, DHS linkage and histone modification patterns employing the 3D genome browser demonstrated that rs4292 regulates the enhancer region of *ACE* in addition to showing long-range chromatin interaction with *FTSJ3* (a gene involved in RNA transcription activity) and *PSMC5* (a gene encoding a protein interacting with Tau)(Figure 4a). rs815460 regulates the enhancer region of *ERC2* and further demonstrates distal chromatin interaction with the enhancer region of *WNT5A* (Figure 4b). *WNT5A* functions in the *wnt* signaling pathway and contributes to Aβ neurotoxicity and neuroinflammation [38]. Analyses of GTEx eQTL data from brain tissue further confirmed that rs4292 is a significant eQTL of *ACE* in the hippocampus (p-value: 7.3E-6) and rs815460 is a significant eQTL of *WNT5A* (p-value: 2.3E-5).

**Figure 4.**
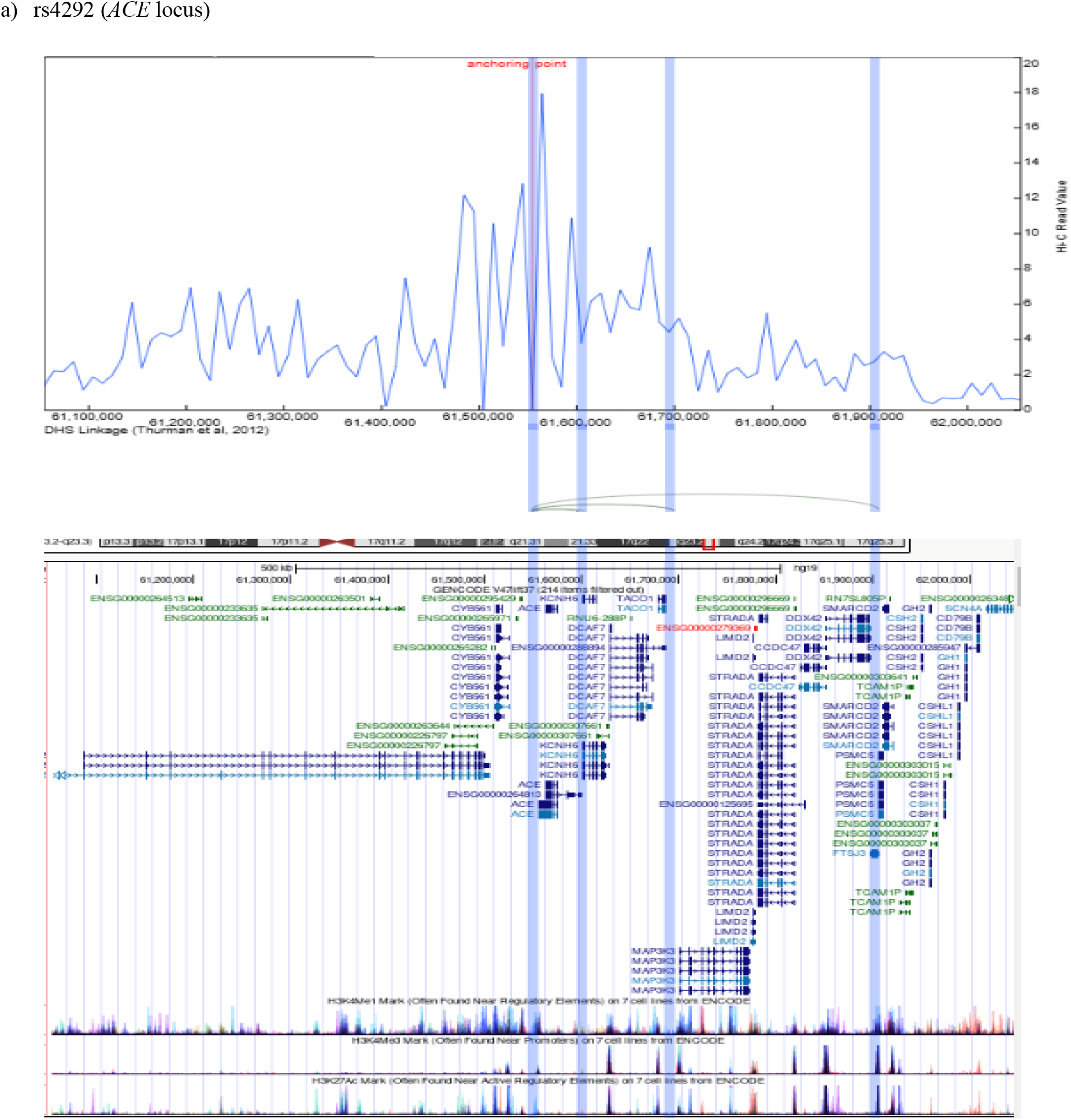
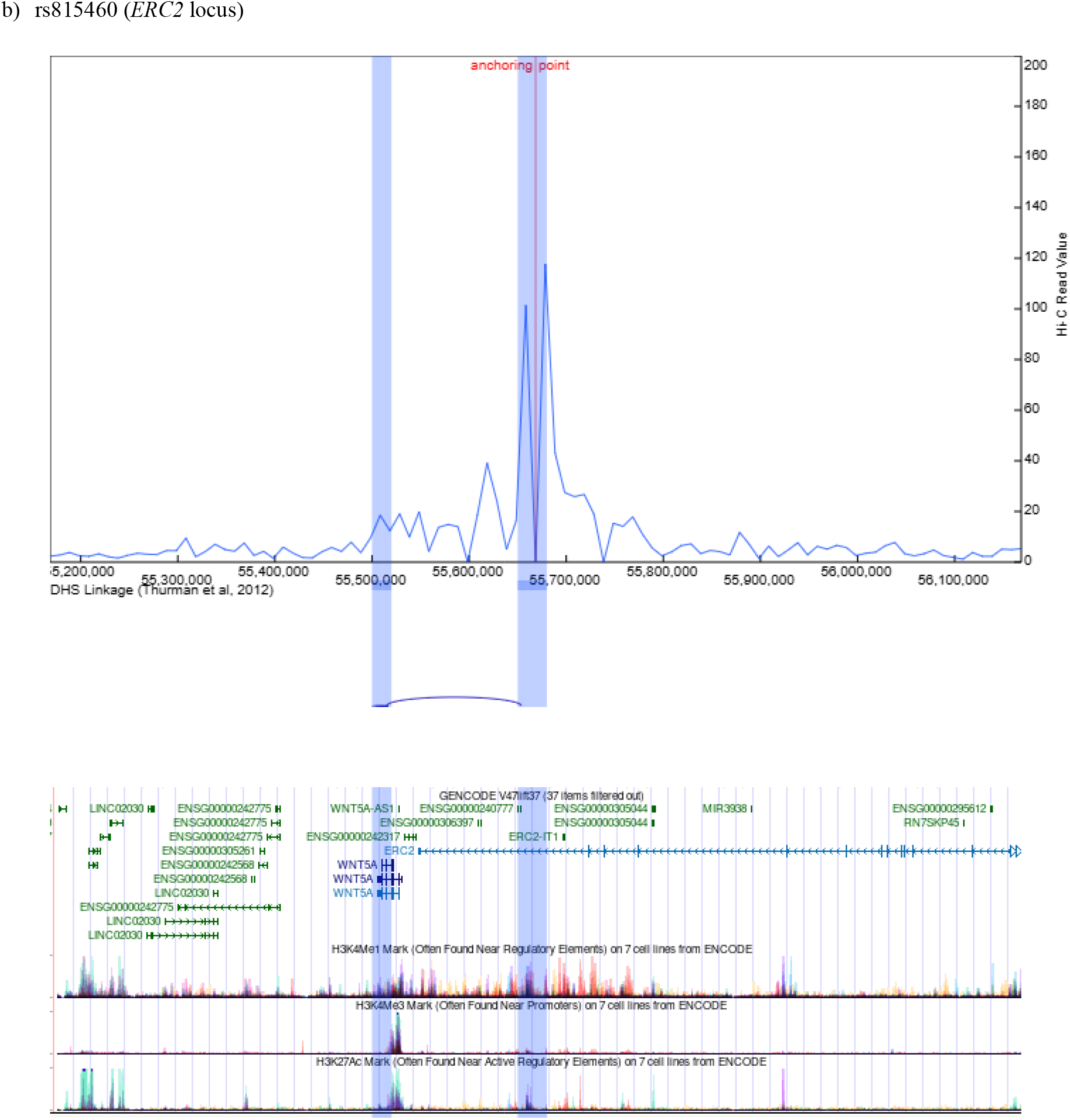
Hi-C analysis of rs4292 (*ACE* locus) and rs815460 (*ERC2* locus); 4C virtual plot with DHS linkage patterns and embedded UCSC genome browser information.

## 4. DISCUSSION

To identify shared genetic loci and mechanistic pathways between AD and primary psychiatric disorders, we conducted global and local genetic covariance analyses between AD and depression, schizophrenia, bipolar disorder, and anxiety, respectively, capitalizing on the largest most recent GWAS summary statistics for each of these traits in non-Hispanic White individuals. Quantification of global polygenic overlap followed by functional finemapping identified both unique genetic components underlying these traits as well as shared polygenic components.

Quantification of global polygenic overlap using MiXeR estimated that roughly 900 independent loci are shared with depression, ∼1,300 independent loci are shared with schizophrenia, ∼1,300 independent loci are shared with bipolar disorder, and ∼1,100 independent loci are shared with anxiety. This suggests that a considerable proportion of variants underlying AD also impacts each of these four psychiatric traits, albeit to different degrees, with 50% of AD variants also being associated with depression but a higher proportion (over 70% of variants) also being associated with schizophrenia and bipolar disorder, and over 60% variants associated with anxiety. While the observed genome-wide polygenic overlap with depression and anxiety showed overall a negative correlation indicating on average opposing effects of causal variants, the shared genetic architectures with both schizophrenia and bipolar disorder presented positive correlations indicating on average the effects of causal variants moved in a similar direction.

Local genetic covariance analyses identified loci at *TMEM106B* on 7p21 as shared between AD and depression as well as anxiety, *ACE* on 17q23 as shared with schizophrenia, and *ERC2* on 3p14 as shared with anxiety. Subsequent fine mapping and sequence data analysis pinpointed a missense variant in *TMEM106B* (rs3173615), a regulatory region variant in *ACE* (rs4292), and two NMD transcript variants in *ERC2* (rs17288728; rs815460) as disease-associated. *TMEM106B* is a ubiquitously expressed type II transmembrane protein localized to the late endosome and lysosome [39, 40] compartments that impacts mass, transport and function of lysosomes. [41] It is associated with brain aging and multiple neurodegenerative diseases, including AD, amyotrophic lateral sclerosis (ALS), frontotemporal lobar degeneration (FTLD), limbic-predominant age-related TAR DNA binding protein 43 (TDP-43) encephalopathy, and Parkinson’s disease [40, 42]. In agreement with this finding, amyloid fibrils containing a *TMEM106B* C-terminal fragment have been observed in brains of individuals with tauopathy, synucleinopathy, Aβ-amyloidosis as well as TDP-43 proteinopathy [43-45]. Genetic studies identified a risk/protective haplotype with several variants in high LD. The major allele (A) of the most widely studied genetic variant in this haplotype, rs1990622, is located in the regulatory sequence downstream of *TMEM106B* and associated with FTLD with TDP-43 inclusions (FTLD-TDP), in particular for individuals with granulin (GRN) gene mutations [46] This allele is associated witha smaller volume of the superior temporal gyrus [47], lower cortical gray matter volumes in the frontal, temporal, cingulate, and insula cortices [48], more advanced TDP-43 pathology at autopsy [49], increased biological aging in the prefrontal cortex [50], worse cognitive function [50], decreased neuronal proportion [51], increased neuronal proportion [52], and *TMEM106B* fibril formation and myelin lipid homeostasis in the aging human hippocampus [53]. Functional finemapping in our analyses followed by analysis of sequence data identified the same haplotype as associated with AD, shared with depression and anxiety, and within this locus missense variant rs3173615 (c.554C□>□G, p.Thr185Ser) - in high LD with rs1990622 - as the most likely causative variant (Supplemental Figure 8).

*ACE* encodes the angiotensin-converting enzyme, a critical component of the renin-angiotensin system (RAS) involved in blood pressure regulation. In recent years, RAS has attracted considerable interest beyond its traditional role in cardiovascular function due to emerging data indicating an involvement in neurodegenerative diseases including AD, and the potential for drug repositioning of RAS modulators to prevent or treat these neurodegenerative disorders [54]. *ACE* converts angiotensin I (Ang I) to angiotensin II, which in turn activates the AT1 receptor (AT1R) resulting in neurotoxicity, neuroinflammation, oxidative stress, beta-amyloid deposition, and hyperphosphorylation of tau protein [54] [55]. While the etiology of schizophrenia is poorly understood, there is evidence that RAS may be involved through direct modulation of inflammation, glutamate, dopamine, GABA, and peroxisome proliferator-activated receptor gamma, all of which are associated with schizophrenia [56]. The regulatory region variant identified by functional finemapping in the present analyses (rs4292) is in high LD (R2=0.93) with a non-coding upstream variant (rs4277405) previously reported by Kunkle et al [55]. However, it is in limited LD (R^2^ = 0.38) with the well-known functional I/D polymorphism rs4646994, localized to intron 16, whose D allele is associated with enhanced *ACE* activity. This indicates that the variant identified in the present analyses likely exerts an effect independent of this indel. In support of this, Hi□C and DHS analyses showed interactions of rs4292 with the promoter region of *ACE* but also long□range looping to *FTSJ3* and *PSMC5. FTSJ3* is an RNA methyltransferase involved in 2′-O-methylation of ribosomal RNA, critical for ribosome biogenesis and protein synthesis, and dysregulation of these processes can impair neuronal function [57]. *PSMC5* is an ATPase component of the 26S proteasome, essential for protein degradation via the ubiquitin-proteasome system crucial for clearing misfolded or toxic proteins including amyloid-β, tau, and α-synuclein [58]. Activation of *PSMC5* (Rpt6) via kinases such as CaMKII has been shown to enhance degradation of α-synuclein in models of Lewy body dementia [58].

*ERC2* (ELKS/RAB6-interacting/CAST family member 2) encodes a presynaptic scaffolding protein essential for organizing the synaptic active zone, where it regulates calcium channel localization and synaptic vesicle docking to support efficient neurotransmitter release. *ERC2* is highly enriched in glutamatergic neurons [59], which are critically involved in excitatory signaling, learning, and memory. Synaptic dysfunction in glutamatergic neurons [59, 60], including impaired NMDA receptor function, is a key contributor to cognitive decline [59, 61]. In line with the observed association with anxiety, altered synaptic structure and plasticity has also been shown to be associated with behavior [60]. Our HiLC and DHS analyses of the two identified NMD transcript variants at the *ERC2* locus showed intra□gene enhancer chromatin interactions.

However, rs815460 also showed distal looping to *WNT5A. WNT5A* encodes a member of the Wnt family of secreted signaling proteins, which play essential roles in regulating cell polarity, migration, synapse formation, and plasticity through non-canonical (β-catenin-independent) signaling pathways. In the central nervous system, *WNT5A* is crucial for proper neuronal development and for maintaining synaptic structure and function, and dysregulation of *WNT5A* signaling has been implicated in neuroinflammation and synaptic degeneration in both AD and psychiatric disorders [38, 62].

This study has several strengths. It is one of the largest studies assessing global and local genetic covariance between AD and major psychiatric disorders. The summary statistics for AD and NPS were obtained from the most recent and largest reported GWAS studies. In addition, to further validate shared identified loci, this study was able to capitalize on a variety of large-scale multi-omics data providing significant supportive evidence for the plausibility of candidate genes at identified loci. A limitation is that we cannot exclude the possibility that we missed additional regions of genetic covariance due to a lack of statistical power, Our findings support the notion of pleiotropic overlap of AD with depression, schizophrenia, bipolar disorders, and anxiety, pinpointing *TMEM106B, ACE*, and *ERC2* as shared genetic drivers. While comprehensive functional characterization of these loci and molecular pathways will be critical to further characterize the specific biological processes through which they exert their effects on these traits, the identification of molecular pathways associated with these loci has important implications for the development of improved therapeutic targets for early treatment and prevention of these disorders.

## Data Availability

This study analyzed third-party GWAS summary statistics. A portion of the data is controlled-access and the remainder is openly available. No individual-level data were used, and this work did not generate new human datasets.
Controlled-access: Million Veteran Program (MVP) GWAS summary statistics obtained via dbGaP (accession phs001672.v1.p). Access is controlled and requires an approved data-access request and data-use agreement.
Openly available:
1. Alzheimer disease: Bellenguez et al. (2022), EBI GWAS Catalog accession GCST90027158 (DOI: 10.1038/s41588-022-01024-z).
2. Bipolar disorder: Mullins et al. (2021), publicly available via the PGC Results & Downloads portal (DOI: 10.1038/s41588-021-00857-4).
3. Schizophrenia: Trubetskoy et al. (2022), publicly available via the PGC schizophrenia downloads page (DOI: 10.1038/s41586-022-04434-5).
4. Anxiety disorders: Friligkou et al. (2024), Zenodo DOI 10.5281/zenodo.13135834.

