## Supplemental Data 1 for "Genetic correlation analysis identifies *TMEM106B, ACE*, and *ERC2* as genetic loci shared between Alzheimer’s disease and primary psychiatric disorders"

**SUPPLEMENTAL MATERIAL**

Table of Contents

Supplementary Table 1. Univariate MiXeR estimates for AD and psychiatric traits.	2

Supplementary Table 2**. Polygenic overlap between Alzheimer’s Disease and neuropsychiatric traits**	3

Supplementary Table 3. Results from local genetic covariance analyses for regions meeting a significance threshold of P <5* 10^-3…………..4,5

Supplementary Table 4. Results from methylation data analyses in the ROSMAP cohort…………………………………………………………..6

Supplementary Figure 1. Univariate Q-Q plots………………………………………………………………….…………………………….……..7

Supplementary Figure 2. Local association plots for genetic loci shared between AD and depression……………..…………………………….....8

Supplementary Figure 3. Local association plots for genetic loci shared between AD and schizophrenia……………...……………......................9

Supplementary Figure 4. Local association plots for genetic loci shared between AD and bipolar disorder…………………………………….....10

Supplementary Figure 5. Local association plots for genetic loci shared between AD and anxiety disorder……………………............................11

Supplementary Figure 6. Expression of *TMEM106B*, *ACE*, *MAPT*, *KANSL1*, *KCNG1* and *ERC2* in brain tissue........………………….…......12,13

Supplementary Figure 7. Association of rs3173615 and rs4292 with proteomic and metabolomic endpoints measured in plasma, CSF and brain tissue (ONTIME)……………………………………………………………………………………………………………………………………14

Supplementary Figure 8. Local association plots for *TMEM106B* indicating localization of top SNPs from the present analysis and variants

previously reported to be associated with AD, FTLD and hippocampal sclerosis…………………………………...………………………………15

No table of contents entries found.

**SUPPLEMENTARY TABLES**

**Supplementary Table 1. Univariate MiXeR estimates for AD and psychiatric traits**. h^2^_SNP_: SNP-based heritability estimate. Polygenicity_90_: The number of causal variants with strongest effects required to explain 90% of SNP-based heritability. AIC and BIC are indices of model fit.

|  | **h2SNP** | | **Polygenicity90** | |  |  |
| --- | --- | --- | --- | --- | --- | --- |
|  | **Mean** | **SD** | **Mean** | **SD** | **AIC** | **BIC** |
| **AD** | 0.08 | 0.27 | 1771.4 | 1353.6 | 116.1 | 108.5 |
| **Depression** | 0.03 | 0.008 | 15250.2 | 7197.9 | 9.2 | 2.0 |
| **Schizophrenia** | 0.44 | 0.06 | 15062.7 | 9668.6 | 19.6 | 12.1 |
| **Bipolar disorder** | 0.08 | 0.02 | 15624.5 | 10125.4 | 6.9 | -0.5 |
| **Anxiety** | 0.02 | 0.006 | 15097.1 | 10457.5 | 1.7 | -4.6 |

Abbreviations: AIC, Akaike information criterion; BIC, Bayesian information criterion; SD, standard deviation

**Supplementary Table 2: Polygenic overlap between Alzheimer’s Disease and neuropsychiatric traits**

| Trait1 | Trait2 | Dice (mean) | rho beta (mean) | rho beta (std) | rg (mean) | fraction concordant within shared (mean) |
| --- | --- | --- | --- | --- | --- | --- |
| AD | Depression | 0.1 | -0.13 | 0.81 | -0.01 | 0.45 |
| AD | Schizophrenia | 0.16 | 0.19 | 0.62 | 0.03 | 0.57 |
| AD | Bipolar | 0.16 | 0.23 | 0.62 | 0.04 | 0.59 |
| AD | Anxiety | 0.14 | -0.05 | 0.76 | 0.02 | 0.44 |

Dice: polygenic overlap; rho beta(mean): genetic correlation for shared causal variants; rho-beta(std): standard deviation of shared causal variant correlation; rg(mean): global genetic correlation coefficient; fraction concordant within shared (mean): proportion of shared causal genetic variants with concordant (same direction) effects

**Supplemental Table 3.** **Results from local genetic covariance analyses for regions meeting a significance threshold of P < 5*10^-3**

| AD vs Depression | | | |
| --- | --- | --- | --- |
| Genomic Regions(hg37) | **Rho** | **R2** | **P < 5*10^-3** |
| chr1:164.7-165.5 Mb | 0.87 | 0.75 | 0.0003 |
| chr2:12.4-12.5 Mb | -0.85 | 0.73 | 0.0008 |
| chr2:23.4-23.5 Mb | 1.00 | 1.00 | 8.03E-05 |
| chr6:1.8-1.9 Mb | 0.97 | 0.95 | 0.0005 |
| chr6:7-7.1 Mb | 1.00 | 1.00 | 0.0004 |
| chr7:11.8-12.6 Mb | -0.80 | 0.64 | 0.0008 |
| chr8:64.2-66 Mb | 0.55 | 0.31 | 0.0005 |
| chr10:87.2-88.4 Mb | 1.00 | 1.00 | 0.0009 |
| chr10:106.1-107.8Mb | 0.71 | 0.51 | 0.0003 |
| chr11:131.424-132.584 Mb | -0.76 | 0.58 | 5.29E-05 |
| chr12:129.887-130.630 Mb | -1.00 | 1.00 | 0.0002 |
| chr13:112.319-113.573 Mb | 0.96 | 0.92 | 0.0008 |
| chr16:5.783-6.446 Mb | 0.66 | 0.43 | 0.0009 |
| chr16:11.023-11.917 Mb | 1.00 | 1.00 | 1.08E-06 |
| chr17:10.573-11.778 Mb | 0.77 | 0.59 | 0.0006 |
| chr20:18.959-19.958 Mb | 0.66 | 0.43 | 0.0006 |
| chr21:26.468-27.774 Mb | 0.58 | 0.34 | 0.0008 |
| AD vs Schizophrenia | | | |
| Genomic Regions(hg37) | **Rho** | **R2** | **P < 5*10^-3** |
| chr1:199.3-200.1 Mb | -1.00 | 1.00 | 0.0010 |
| chr2:1.9-2.7 Mb | 0.82 | 0.67 | 9.02E-06 |
| chr3:116.5-117.2 Mb | -1.00 | 1.00 | 0.0006 |
| chr3:123.5-124.8 Mb | -1.00 | 1.00 | 0.0009 |
| chr3:124.8-125.5 Mb | 0.78 | 0.60 | 0.0008 |
| chr4:168.6-169.6 Mb | 1.00 | 1.00 | 0.0004 |
| chr6:112.3-113.7 Mb | -1.00 | 1.00 | 0.0001 |
| chr7:137.7-138.8 Mb | -0.64 | 0.42 | 8.21E-05 |
| chr8:52.3-53.1 Mb | -0.91 | 0.82 | 0.0003 |
| chr9:128.8-129.6 Mb | 0.61 | 0.38 | 0.0003 |
| chr9:138.2-139.0 Mb | -0.71 | 0.50 | 0.0004 |
| chr10:12.6-13.2 Mb | 0.80 | 0.64 | 0.0005 |
| chr11:124.4-125.3 Mb | 0.51 | 0.26 | 0.0001 |
| chr12:57.0-58.7 Mb | 0.88 | 0.77 | 9.49E-05 |
| chr13:112.3-113.6 Mb | 0.83 | 0.69 | 0.0003 |
| chr3:116.5-117.2 Mb | 0.70 | 0.49 | 0.0006 |
| chr17:43.5-44.9 Mb | 0.73 | 0.53 | 1.14E-05 |
| chr17:60.5-62.1 Mb | 0.79 | 0.63 | 8.45E-06 |
| chr18:52.5-53.8 Mb | 0.81 | 0.66 | 5.14E-07 |
| chr18:72.8-73.6 Mb | 0.83 | 0.69 | 0.0003 |
| chr19:48.6-49.4 Mb | 0.92 | 0.85 | 0.0010 |
| chr20:49.2-50.7 Mb | 0.72 | 0.52 | 3.35E-08 |
| chr20:62.2-63.0 Mb | 0.74 | 0.54 | 0.0008 |
| AD vs Bipolar disorder | | | |
| Genomic Regions(hg37) | **Rho** | **R2** | **P < 5*10^-3** |
| chr3:36.8-38.7 Mb | 0.64 | 0.41 | 2.81E-05 |
| chr4:86.9-88.5 Mb | 1.00 | 1.00 | 0.0007 |
| chr6:112.3-113.7 Mb | -0.95 | 0.90 | 0.0008 |
| chr6:1238.5-1253.7 Mb | 0.73 | 0.53 | 0.0009 |
| chr6:127.5-128.8 Mb | 0.73 | 0.54 | 0.0008 |
| chr6:148.2-149.3 Mb | 0.87 | 0.75 | 0.0005 |
| chr7:31.7-32.5 Mb | 1.00 | 1.00 | 0.0003 |
| chr8:28.3-29.7 Mb | -1.00 | 1.00 | 0.0003 |
| chr8:137.5-138.6 Mb | 0.83 | 0.69 | 0.0004 |
| chr11:27.0-28.6 Mb | -1.00 | 1.00 | 0.0003 |
| chr11:130.5-131.4 Mb | -0.91 | 0.83 | 2.38E-05 |
| chr12:53.0-54.4 Mb | 0.93 | 0.86 | 0.0009 |
| chr17:65.6-66.8 Mb | 0.64 | 0.41 | 0.0003 |
| chr18:59.4-60.8 Mb | -1.00 | 1.00 | 0.0001 |
| chr19:6.5-7.2 Mb | -0.93 | 0.87 | 0.0002 |
| chr20:49.2-50.7 Mb | 0.53 | 0.28 | 0.0004 |
| chr22:23.9-25.3 Mb | -0.68 | 0.47 | 0.0006 |
| AD vs Anxiety | | | |
| Genomic Regions(hg37) | **Rho** | **R2** | **P < 5*10^-3** |
| chr2:79.9-80.7 Mb | -0.62 | 0.38 | 0.003 |
| chr3:55.6-56.9 Mb | 0.51 | 0.26 | 0.005 |
| chr6:31.3-31.4 Mb | 0.74 | 0.55 | 0.001 |
| chr7:11.8-12.6 Mb | -0.77 | 0.59 | 0.003 |
| chr7:17.5-19.1 Mb | -0.79 | 0.63 | 0.002 |
| chr11:46.3-48 Mb | 0.84 | 0.72 | 0.001 |
| chr14:33.5-34.6 Mb | -0.57 | 0.33 | 0.005 |
| chr15:90.6-91.5 Mb | 0.76 | 0.57 | 0.003 |
| chr15:96.8-98 Mb | -0.66 | 0.44 | 8.52E-05 |

Rho: correlation coefficient between the two traits; r2: proportion of genetic factors shared between the two traits; P<0.005: p-value threshold for significant association

**Supplemental Table 4. Results from methylation data analyses in the ROSMAP cohort**

| **Gene** | **TargetID** | **tangles coefficient** | **tangles p-value** | **amyloid coefficient** | **amyloid p-value** |
| --- | --- | --- | --- | --- | --- |
| *TMEM106B* | cg25424200 | 0.000247952 | 4.77E-01 | -0.00139308 | **1.41E-03** |
| *ACE* | cg06751221 | -0.00062033 | 2.07E-01 | 0.00184962 | **2.61E-03** |
| *ERC2* | cg15075580 | -0.000551262 | 6.98E-01 | 0.006020759 | **1.14E-03** |

Models are adjusted for age at death, sex, experimental batch, and bisulfite conversion efficiency

**Supplementary Figure 1. Univariate Q-Q plots for distribution of expected p-values under a null model (no SNPs associated with the phenotype) (x axis) versus observed p-values (y axis).** Univariate Q-Q plots demonstrate that MiXeR-based predictions provide accurate estimates of the data Q-Q plots. Blue lines indicate p-values of SNPs observed in GWAS summary statistics with grey shading indicating 95% confidence interval. Orange lines indicate model predictions. The dashed line is the expected Q-Q plot under null (no SNPs associated with the phenotype). The vertical axes are limited to the genome-wide significance threshold of p<5×10−8 to highlight behavior of polygenic component. Points on the Q-Q plot are weighted according to LD structure, using n=64 iterations of random pruning at LD threshold r2=0.1.

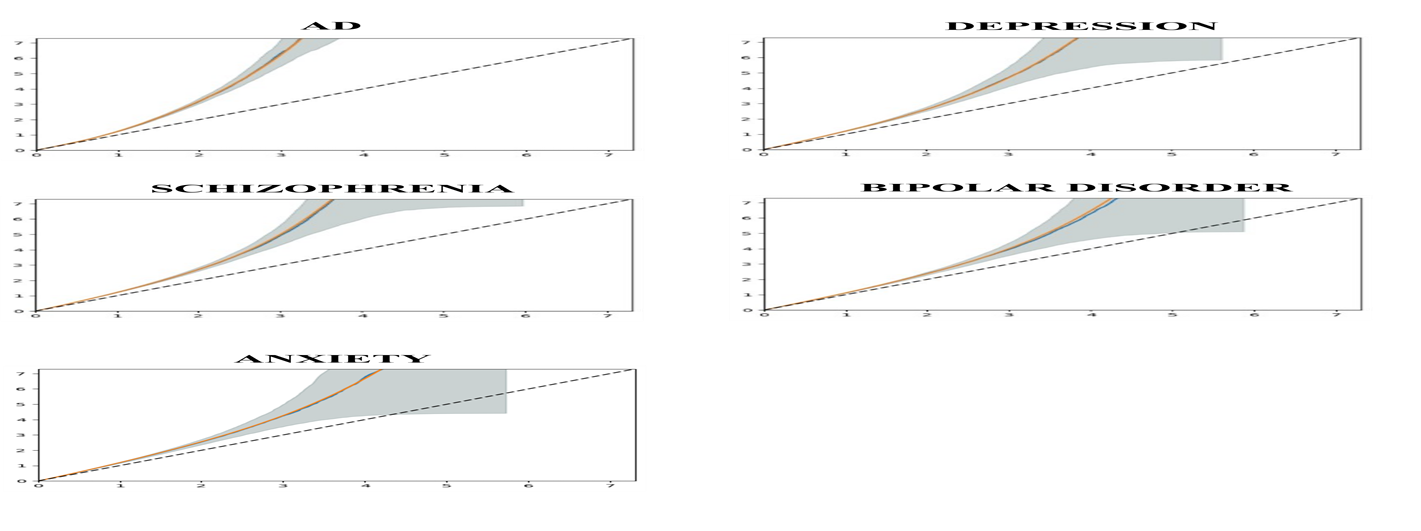

**Supplementary Figure 2.** **Local association plots for genetic loci shared between AD and depression.** The x-axis represents the base pair position on the chromosome. The left y-axis shows -log10(p-values) for genetic association for each genetic variant in the analyses, while the right y-axis depicts recombination rates (cM/Mb). The color of the points indicates the linkage disequilibrium (r²) with the lead SNP.

a) chr2:234.9 -235.6 Mb

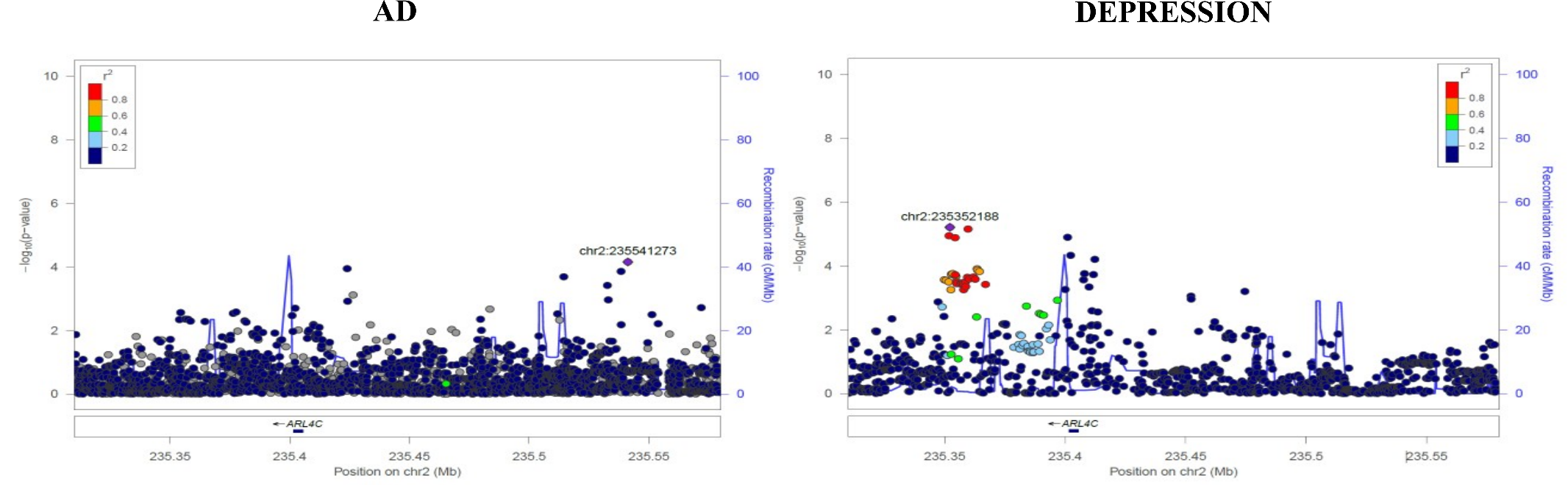

b) chr6:18.7 -19.4 Mb

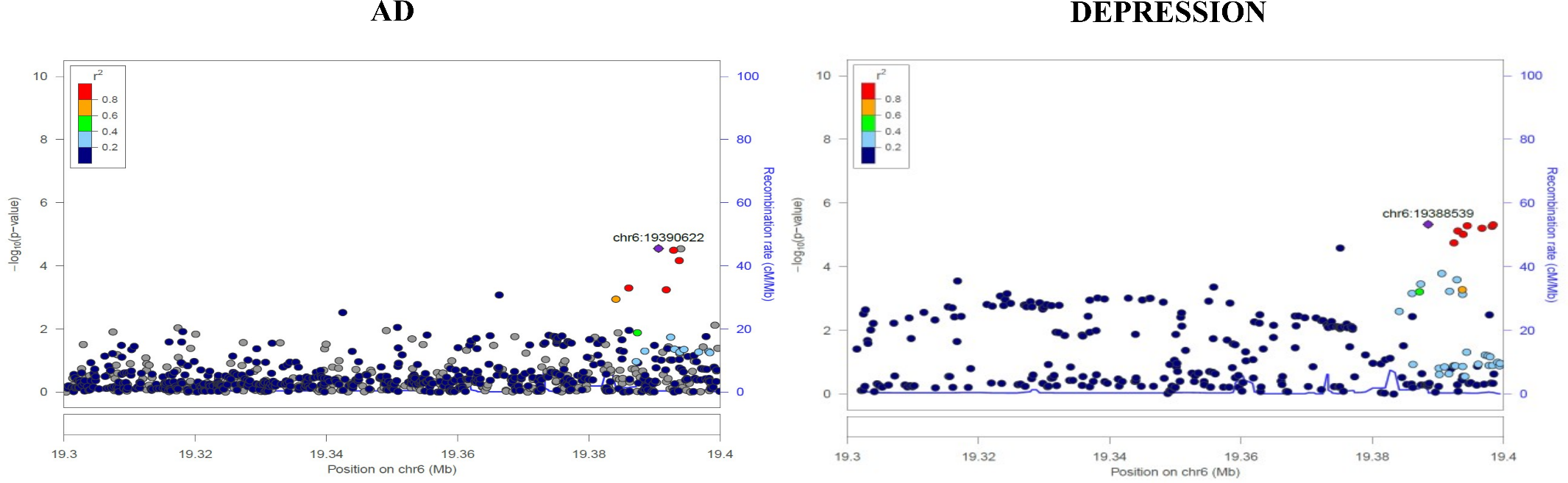
c) chr16:5.7 - 6.4 Mb

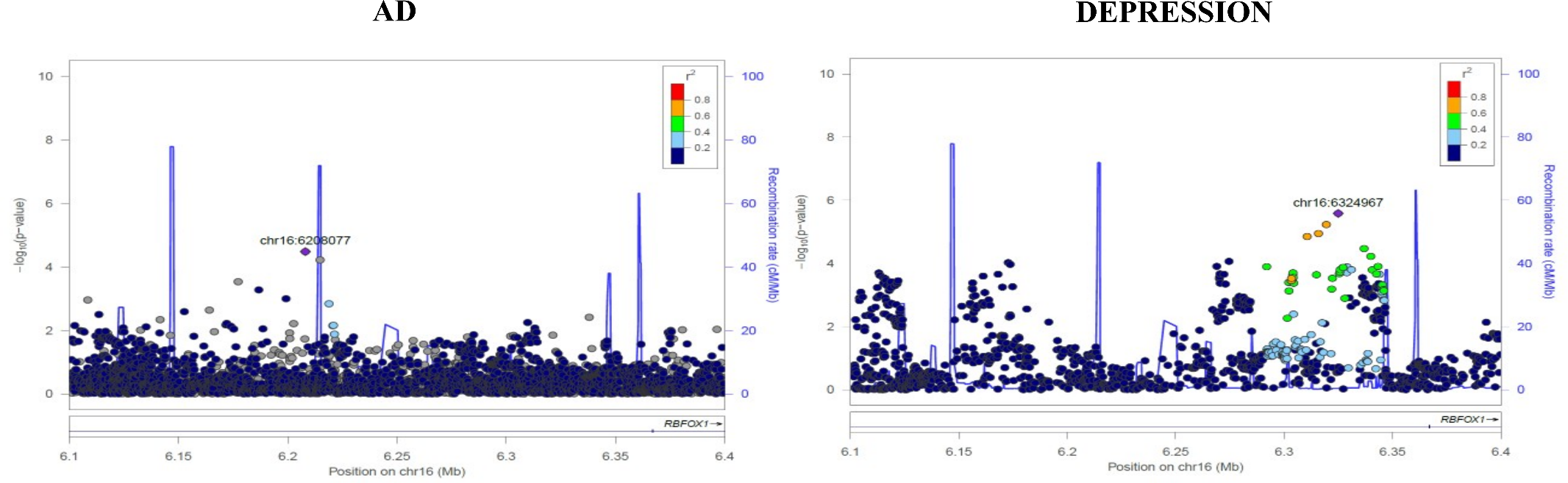

**Supplementary Figure 3.** **Local association plots for genetic loci shared between AD and schizophrenia.** The x-axis represents the base pair position on the chromosome. The left y-axis shows -log10(p-values) for genetic association, while the right y-axis depicts recombination rates (cM/Mb). The color of the points indicates the linkage disequilibrium (r²) with the lead SNP. The left-hand plots represent AD, and the right-hand plots correspond to Schizophrenia.

a) chr3:116.4 -117.2 Mb

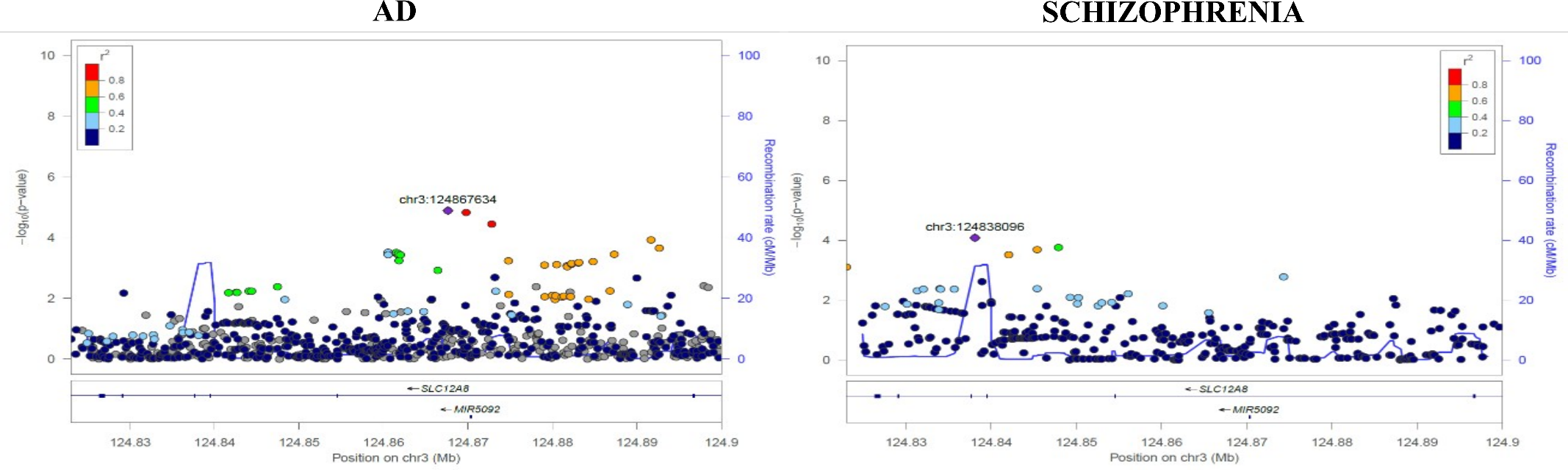

b) chr20:62.1 -62.9 Mb

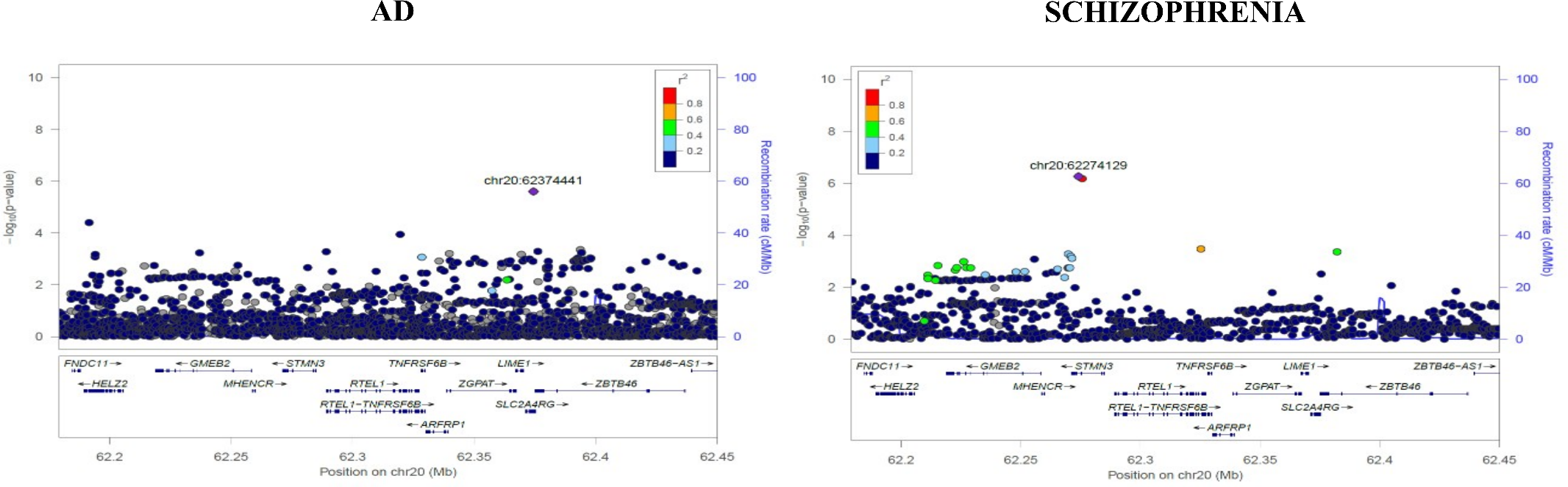

**Supplementary Figure 4.** **Local association plots for identified genetic loci between AD and bipolar disorder.** The x-axis represents the base pair position on the chromosome. The left y-axis shows -log10(p-values) for genetic association, while the right y-axis depicts recombination rates (cM/Mb). The color of the points indicates the linkage disequilibrium (r²) with the lead SNP. The left-hand plots represent AD, and the right-hand plots correspond to bipolar disorder..

a) chr3:36.8-38.7 Mb

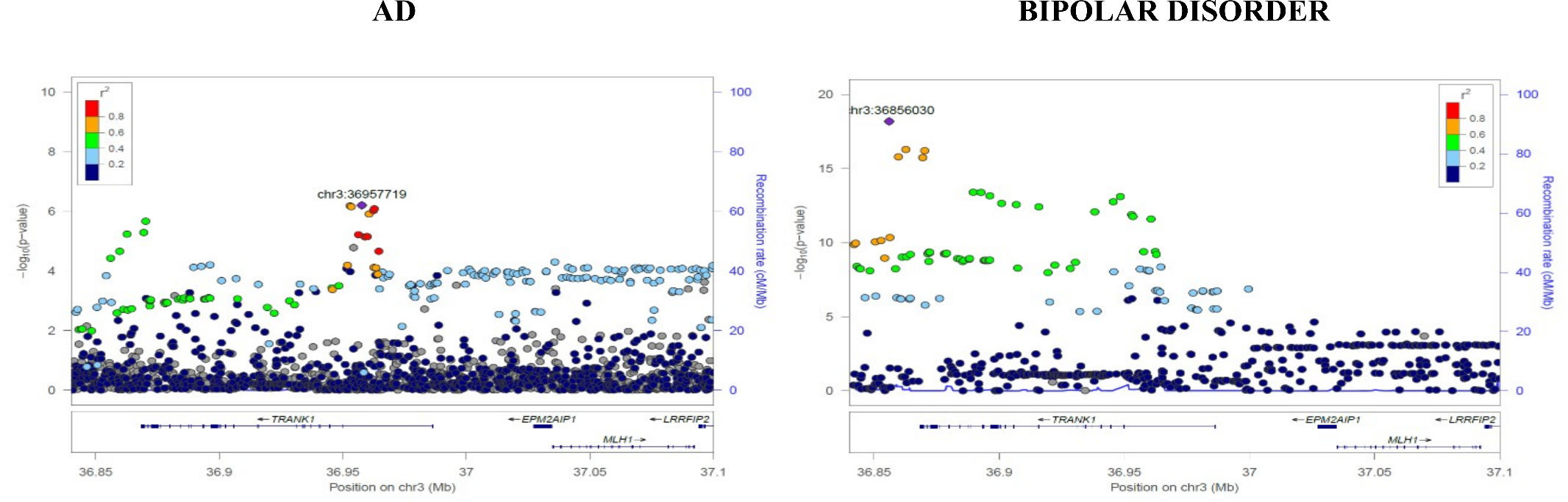

b) chr18:59.4 -60.7 Mb

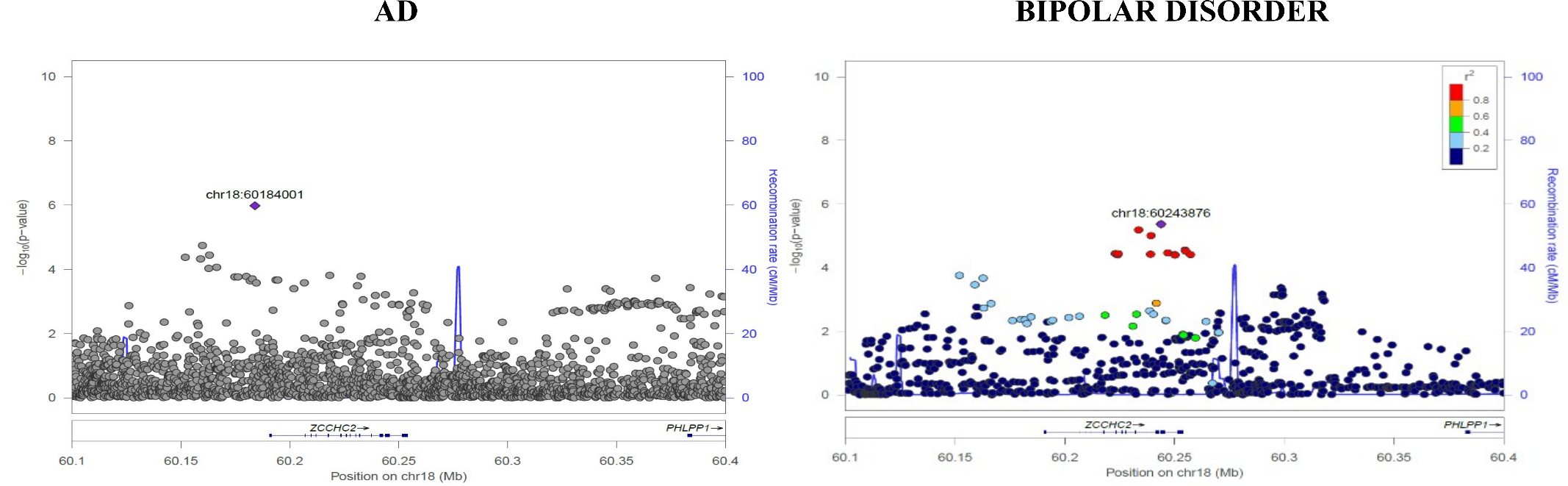

**Supplementary Figure 5.** **Local association plots for genetic loci shared between AD and anxiety.** The x-axis represents the base pair position on the chromosome. The left y-axis shows -log10(p-values) for genetic association, while the right y-axis depicts recombination rates (cM/Mb). The color of the points indicates the linkage disequilibrium (r²) with the lead SNP. The left-hand plots represent AD, and the right-hand plots correspond to Anxiety.

a) chr6:31.3-31.4 Mb

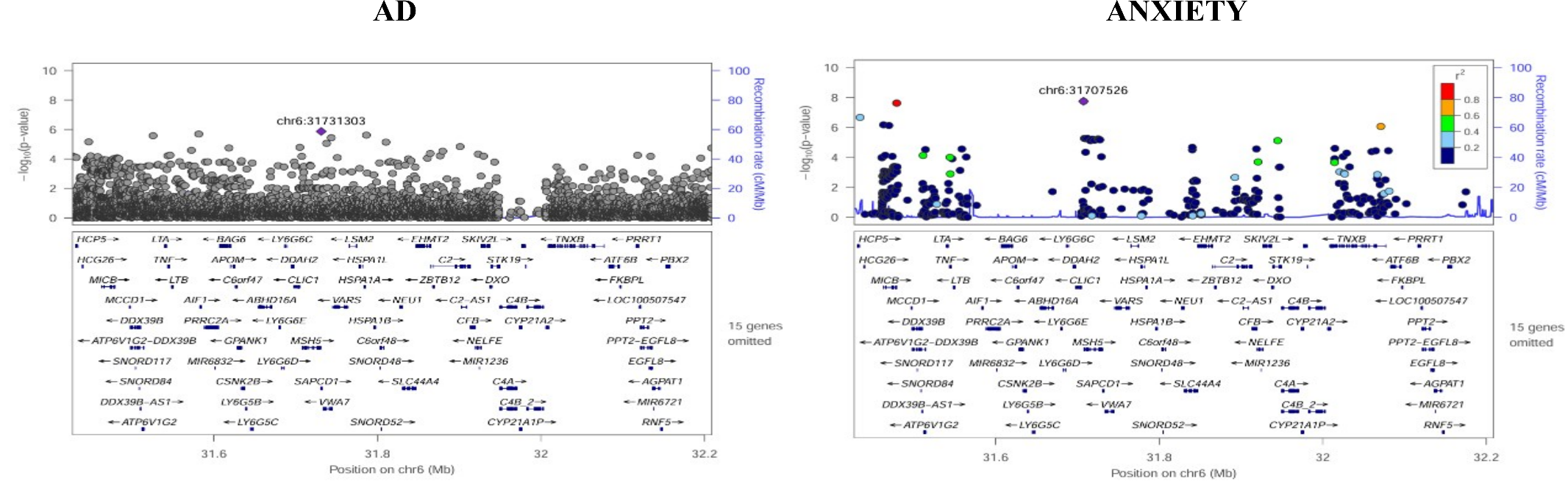

**Supplementary Figure 6. Expression of *TMEM106B, ACE, MAPT, KANSL1, KCNG1* and *ERC2* in brain tissue.** The x-axis shows different brain regions. The y-axis shows LOG2CPM values for gene expression. ACC; Anterior cingulate cortex, CBE; Cerebellum, DLPFC; Dorsolateral prefrontal cortex, FP; Frontal Pole, IFG; Interior frontal gyrus, PCC; Posterior cingulate cortex, PHG; Parahippocampal gyrus, STG; Superior temporal gyrus, TCX; Temporal cortex

a) *TMEM106B*

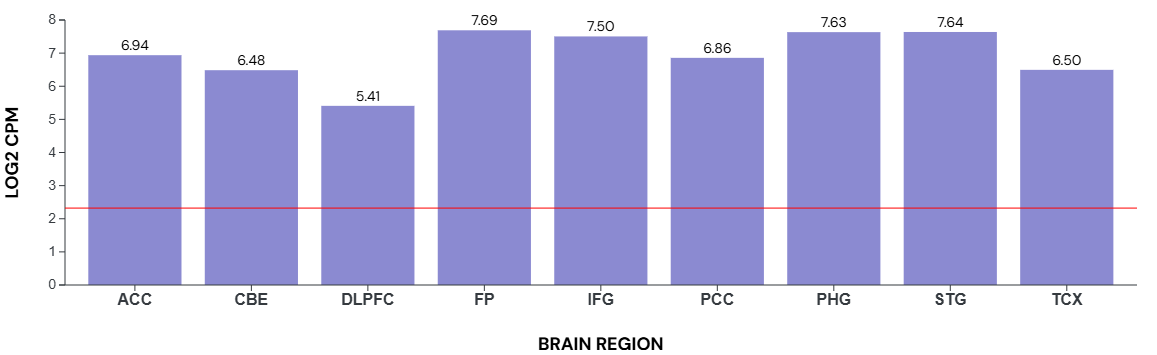

b) *ACE*

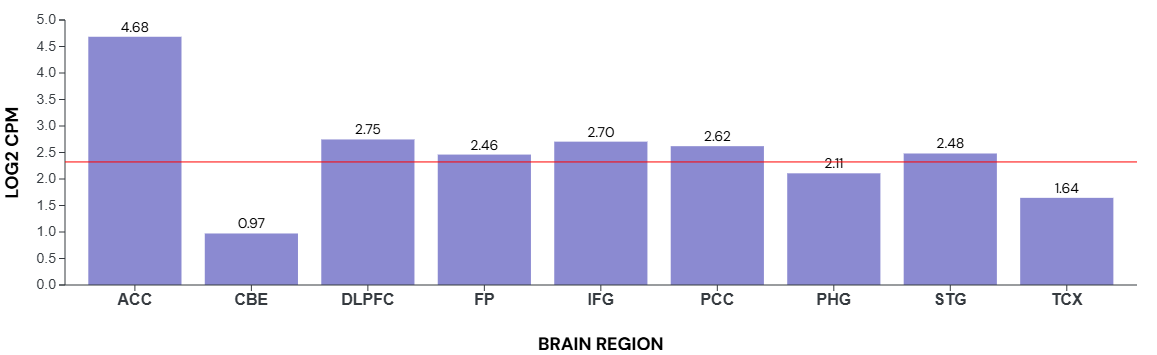

c) *MAPT*

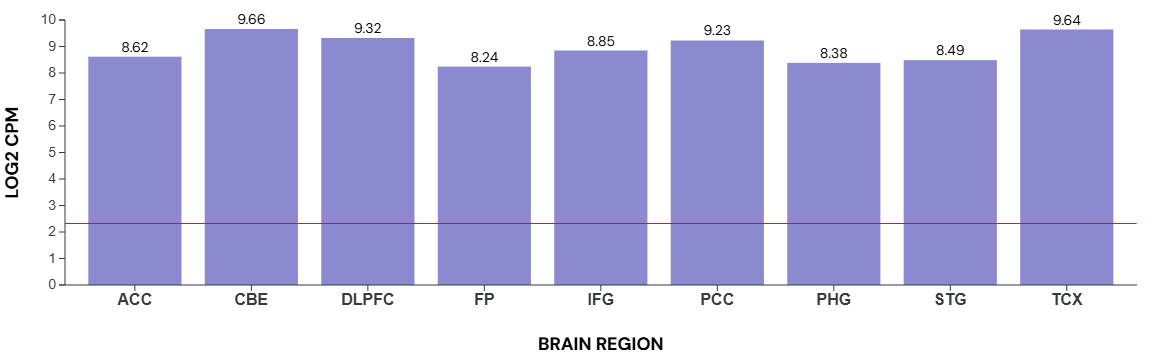

d) *KANSL1*

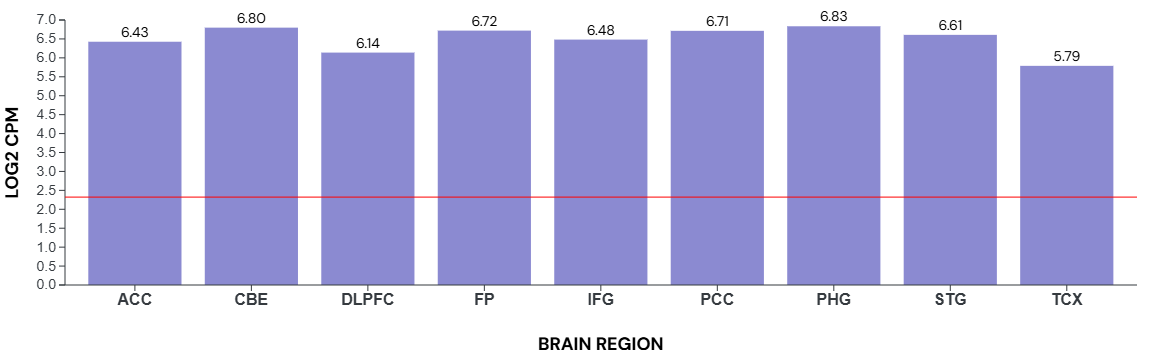

e) *KCNG1*

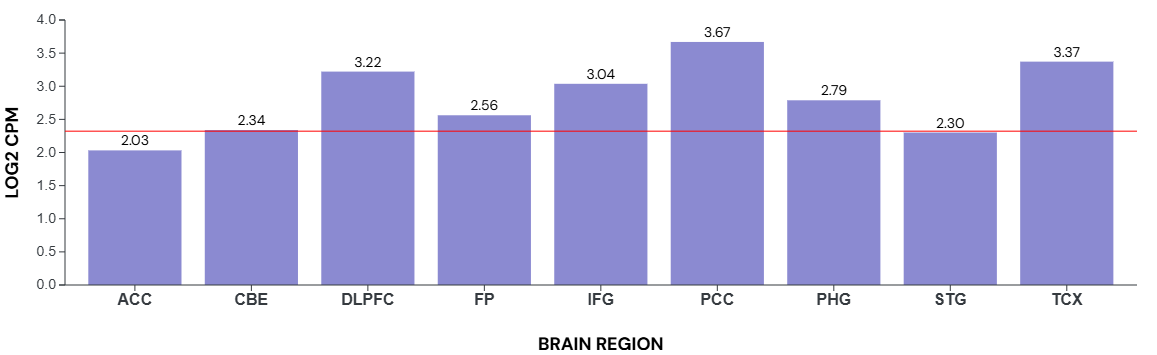

f) *ERC2*

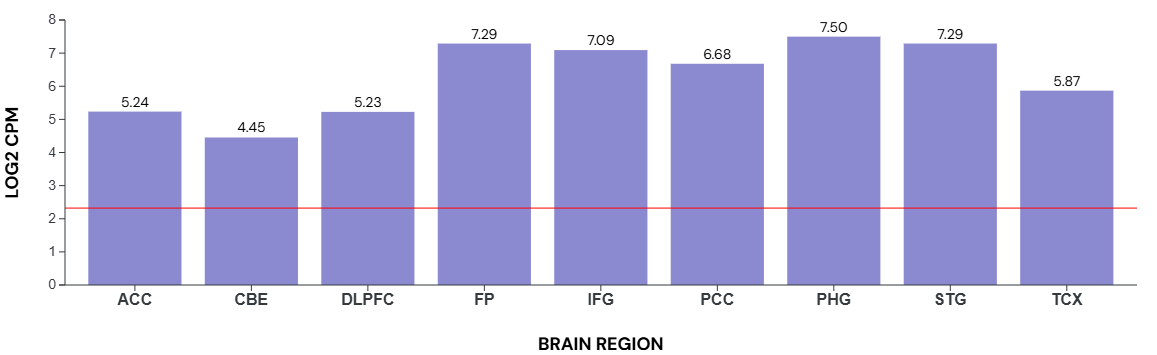

**Supplementary Figure 7. Association of rs3173615 and rs4292 with proteomic and metabolomic endpoints measured in plasma, CSF and brain tissue.** The x-axis shows the various types of multiomics data; the y-axis shows -log10(p-values) for association with identified top variants.

a) rs3173615 (*TMEM106B* missense variant)

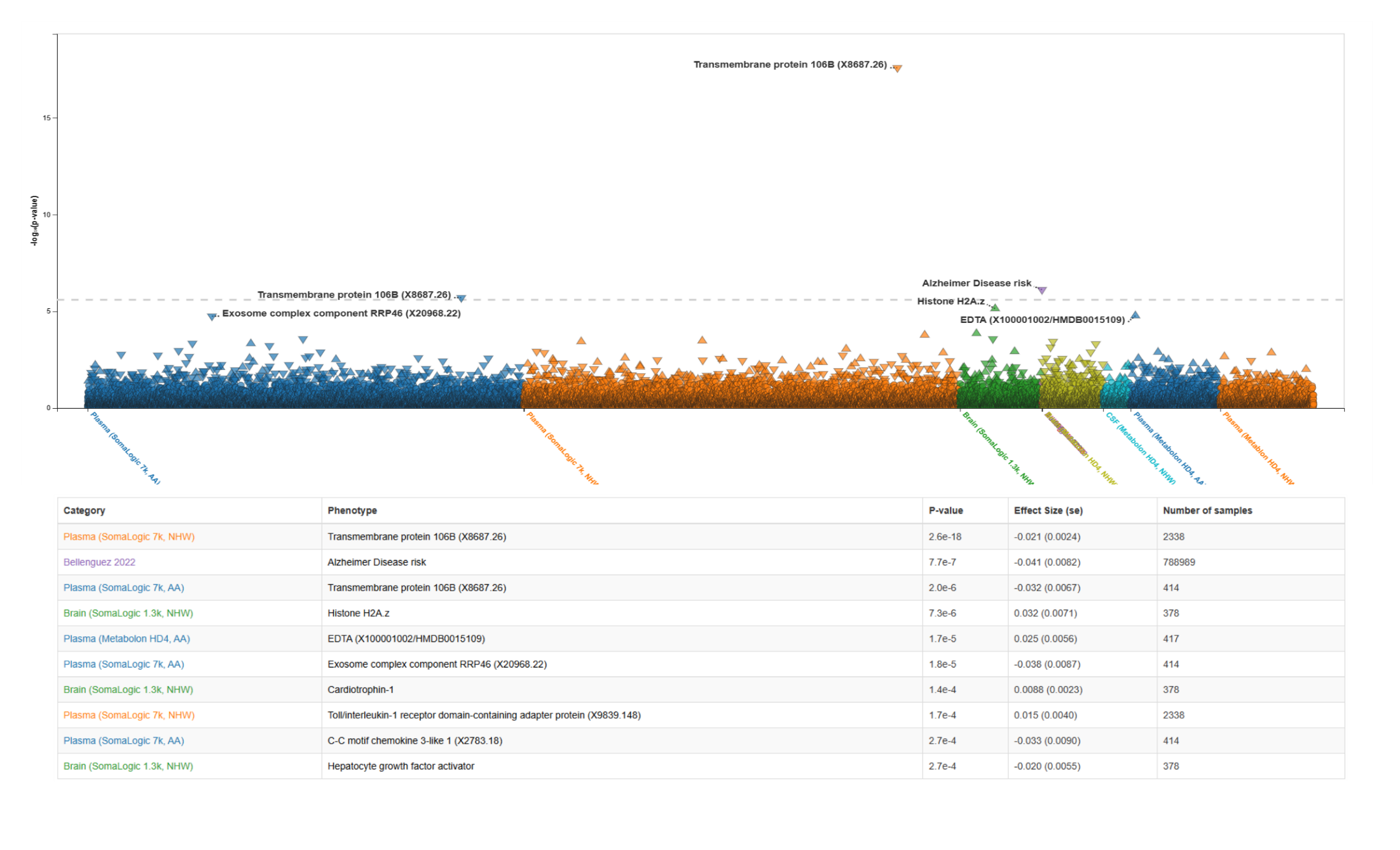

b) rs4292 (*ACE* regulatory region variant)
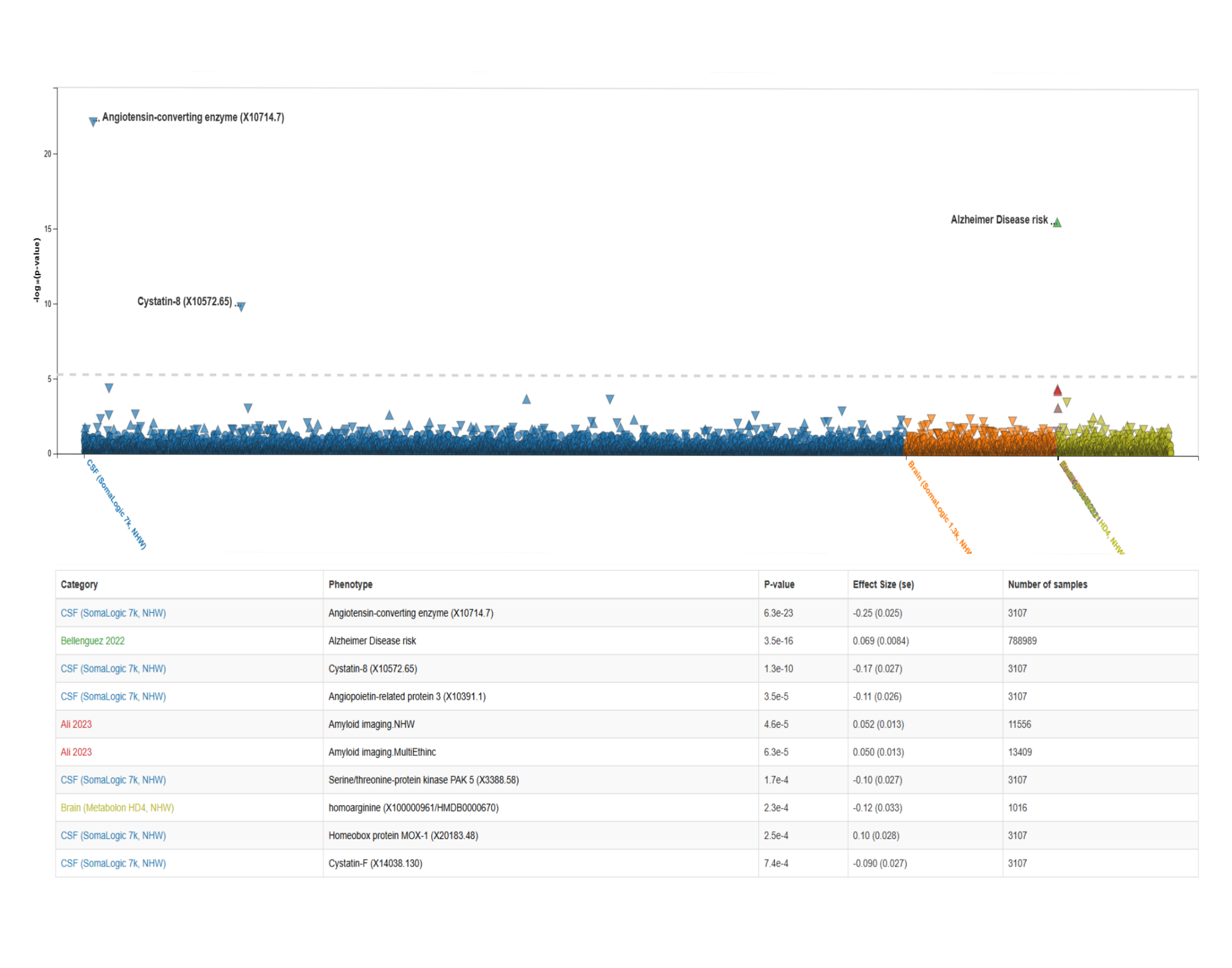

**Supplementary Figure 8.** **Local association plots for *TMEM106B* indicating localization of AD, depression and anxiety top SNPs from the present analysis along with localization of top genetic variants previously reported to be associated with AD, FTLD and hippocampal sclerosis.** The x-axis represents the base pair position on the chromosome. The left and y-axis shows -log10(p-values) for genetic association. The color of the points indicates the linkage disequilibrium (r²) with the GWAS lead SNP for each trait.

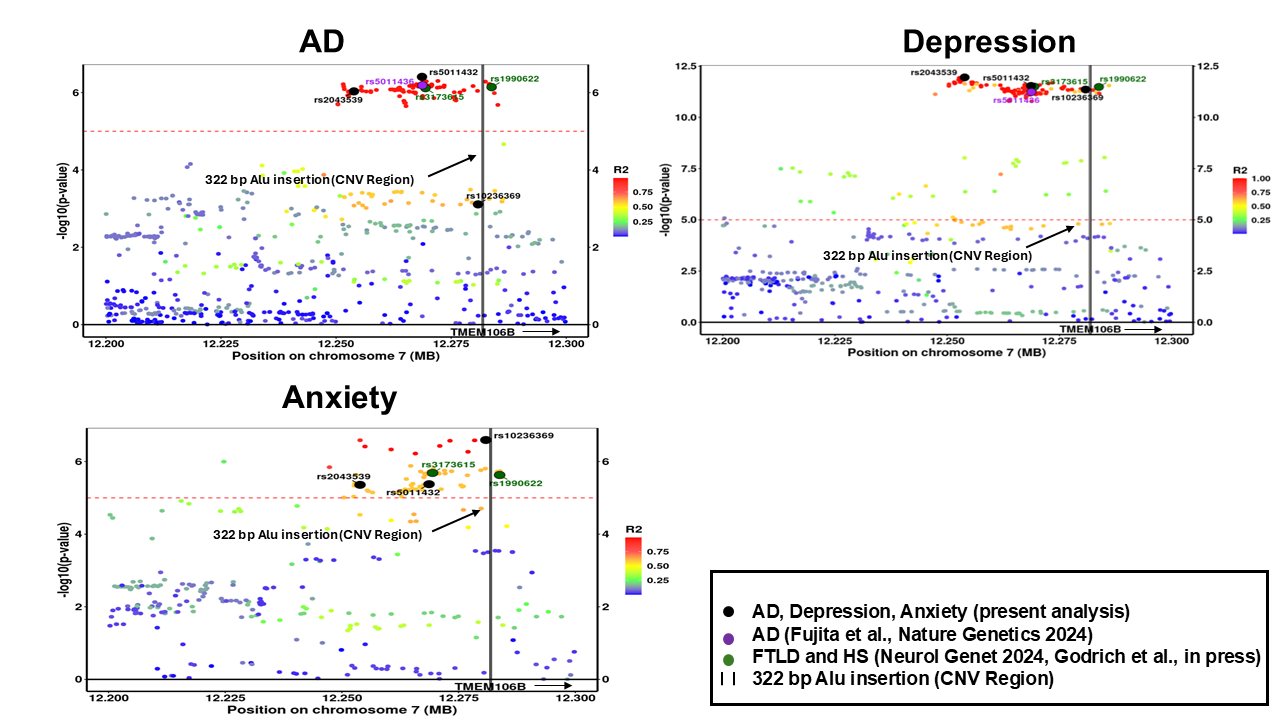
